# Vaccination is reasonably effective in limiting the spread of COVID-19 infections, hospitalizations and deaths with COVID-19

**DOI:** 10.1101/2021.12.12.21267672

**Authors:** Jože P. Damijan, Sandra Damijan, Črt Kostevc

## Abstract

This paper uses large cross-country data for 110 countries to examine the effectiveness of COVID vaccination coverage. Our results confirm that vaccines are reasonably effective in both limiting the spread of infections and containing more severe disease progression in symptomatic patients. First, the results show that full vaccination rate is consistently negatively correlated with the number of new COVID cases, whereby a 10 percent increase in vaccination rate is associated with a 1.3 to 1.7 percent decrease in new COVID cases. Second, the magnitude of vaccination is shown to contribute significantly to moderating severe disease progression. On average, a 10 percent increase in the rate of vaccination leads to a reduction of about 5 percent in the number of new hospitalizations, 12 percent decrease in the number of new intensive care patients and 2 percent reduction in the number of new deaths. Finally, by comparing the data for the same period between 2020 and 2021, we also check how good is vaccination as a substitute for lockdowns or other stringent government protection measures. Results suggest that vaccination does not appear to be an effective substitute for more stringent government safety measures to contain the spread of COVID infections until a high vaccination coverage threshold (more than 70 percent) has been achieved. On the other hand, vaccination is shown to be quite effective in limiting the more severe course of the disease in symptomatic patients already at moderate vaccination coverage (between 40 and 70 percent). This suggests that vaccination can also help to reduce pressure on the health system and thus benefit the overall public health of society.

**Summary:** *Background:* Using a simple descriptive analysis, in a recent paper Subramanian and Kumar (2021) claim that the increase in COVID-19 cases is not related to levels of vaccination across 68 countries and US counties. We demonstrate that this type of analysis, based on only one data point per country and without rigorous analysis of the dynamics of the pandemic and vaccination, is far too simplistic and inevitably leads to false conclusions that are not justified by the data. Using large cross-country data for 110 countries, we estimate two complex models to examine the impact of vaccination coverage on the spread of COVID infections and on the course of severe COVID disease.

*Methods:* We use daily COVID-related data for 110 countries (from Our World in Data) for the period from August 1, 2021 onwards to estimate two comprehensive models – one for the impact on the spread of COVID infections (in terms of number of new confirmed cases) and another one for the impact on severe COVID disease progression (in terms of number of new hospitalizations, admissions to intensive care and deaths). In estimating the vaccines effectiveness, the models capture the differences across countries regarding the state of epidemics and its dynamics, the timing of full vaccination, and country-specific factors. The latter account for differences between countries that determine countries’ specific vulnerabilities or strengths in response to the pandemic. Our data are structured as panel data with a cross-sectional (country) and time (week) dimension. Both models are estimated using a pooled ordinary least squares (OLS) estimator. We first estimate our baseline models and then proceed with two alternative model specifications to examine to what extent vaccination can serve as a substitute for more stringent government protective measures.

*Findings:* Our results confirm that vaccines are reasonably effective in both limiting the spread of infections and containing more severe disease progression in symptomatic patients. First, we show that full vaccination rate is consistently negatively correlated with the number of new COVID cases, whereby a 10 percent increase in vaccination rate is associated with a 1.3 to 1.7 percent decrease in new COVID cases. Second, the estimations show that the magnitude of vaccination contributes significantly to moderating severe disease progression. On average, a 10 percent increase in the rate of vaccination leads to about a 5 percent reduction in the number of new hospitalizations, 12 percent decrease in the number of new intensive care patients and 2 percent reduction in the number of new deaths associated with COVID. Third, the estimations confirm that the moderating effect of vaccines on the number of cases and deaths occurs when the vaccination rate is sufficiently high. Finally, by comparing the data for the same period between 2020 and 2021, we also check how good a substitute vaccination is for lockdowns or less stringent government protection measures. Our results suggest that vaccination does not appear to be an effective substitute for more stringent government safety measures to contain the spread of COVID infections until a high vaccination coverage threshold (more than 70 percent) has been achieved. On the other hand, vaccination is shown to be quite effective in limiting the more severe course of the disease in symptomatic patients already at moderate vaccination coverage (between 40 and 70 percent).

*Interpretation:* Our results show that vaccines are effective in both limiting the spread of infection and containing a more severe course of disease in symptomatic patients. High vaccination coverage has been shown to be a reasonably effective tool and may serve in part as a substitute for more stringent government protective measures. In this way, it can also help to reduce pressure on the health system and thus benefit the overall public health of society during such severe pandemics.

*Funding:* European Union Horizon 2020 grant (GROWINPRO, Grant Agreement No. 822781)

## 1. Introduction

Apart from longitudinal medical observational cohort studies,^8^ there are few cross-country empirical studies that use daily country-level COVID-related data to analyze the effectiveness of vaccination levels across countries on the dynamics of the number of new COVID cases, new hospitalizations, new ICU patients, and new deaths with COVID. To our knowledge, there is only one econometric study that uses cross-country data to analyze the effectiveness of vaccines. Aizenman et al (2021) study the impact of vaccination on the ratio of mortality to infections. On the other hand, there is a widely shared paper that uses descriptive analysis to examine the effectiveness of vaccines on the incidence of new COVID -19 (hereafter: COVID) cases (see Subramanian and Kumar, 2021). However, in spite of its broad popular appeal, this study is fraught with many methodological problems.

In examining the relationship between vaccination rates and the incidence of new COVID cases, Subramanian and Kumar (2021) use a simple static framework. First, they employ bivariate analysis by plotting a graph with the COVID cases per 1 million people for 68 countries on the vertical axis against the latest available percentage of the population that is fully vaccinated on the horizontal axis. No rigorous regression analysis was performed, only correlations were visually assessed to determine that there is no relationship between the two variables. Second, they choose only one cross-sectional observation per country (the last available) to show the relationship between the number of new cases and the vaccination rate. And third, all other factors except vaccination rate that might influence the dynamics of epidemics are ignored.

Let us first show why this approach is misleading and why it gives rise to potentially wrong conclusions. We illustrate two main concerns with this approach. The first issue is related to the use of only one cross-sectional observation per country. To illustrate this, let us take two extreme cases in terms of vaccination rates - Bulgaria and Denmark. As shown in Figure 1, Bulgaria is characterized by very low vaccination rates - between 15 and 22 percent in the period from August 1 to October 31, 2021, while at the same time it was affected by a dramatic increase in new COVID cases (see the steep blue trend line in the left part of the Figure 1). On the other hand, Denmark managed to vaccinate a large portion of its population - between August 1 and October 31, 2021, vaccination rates increased from 55 percent to more than 75 percent. At the same time, new COVID infections did increase, but at a very slow rate (see the flat blue trend line in the right part of the Figure 1).

**Figure 1:**
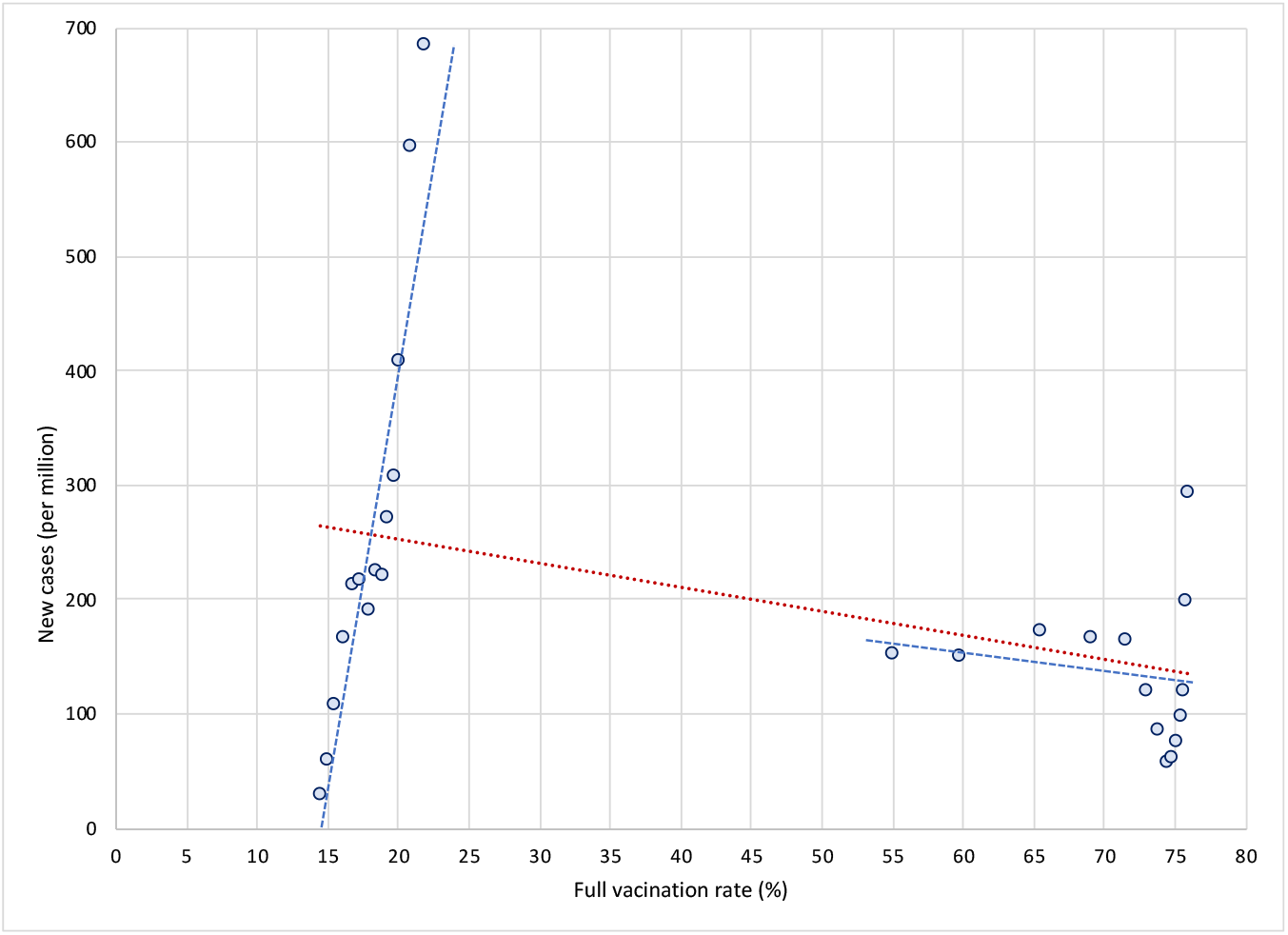
New Covid cases and vaccination rates in Bulgaria and Denmark, by weeks *Notes:* Data by weeks, period August 1 to October 31, 2021. Left part of the figure shows data for Bulgaria *Source*: Our World in Data; own calculations.

This suggests that the high (and increasing) vaccination rates might have dampened the spread of infections in Denmark as opposed to Bulgaria, as shown by a negative (red) trend line for the sample of both countries. In other words, differences in vaccination rates (and in their dynamics in terms of the roll out of vaccines) between countries can have a crucial impact on the dynamics of COVID epidemics over time. High (and increasing) vaccination rates may lead to reduced rates of increase in COVID infections. This can only be observed over a longer time period and in a large sample of countries. If we were to use only one observation per country, as Subramanian and Kumar (2021) have done, we would miss this fundamental insight on the dynamic relationship between vaccination and spread of infections. For this reason, an analysis based on single cross-sectional observation per country is misleading and easily leads to false conclusions that are not justified by the data.

The second issue is related to omitting all factors other than vaccination rates that might influence the dynamics of the pandemic from the analysis. This leads to what is commonly known as the omitted variable problem known in statistics. It occurs when one or more relevant variables are omitted from the empirical model, leading to bias in the results because the model attributes the effect of the missing variables to those that were included. In the economics literature, the problem was popularised, among others, by Barro (1991), who used a sample of 98 countries over the period 1960-1985 to try to uncover the factors that drive economic growth. Estimating the relationship between the growth rate of real GDP per capita and the initial (1960) level of real GDP per capita in a bivariate setting, Barro finds a positive but insignificant relationship between the two. Thus, based on this one might conclude that there is no empirical support for the Solow-Swan hypothesis of convergence across countries. However, after including additional factors in the model (such as differences in human capital, fertility rates, ratio of physical investment to GDP, share of government consumption to GDP, share of public investment, measures of political stability, and a proxy for market distortions) Barro finds that the relationship between the growth rate of real GDP per capita and the initial level of real GDP per capita turns to negative and significant. This means that when important factors are omitted, the model may attribute the effect of the missing variables to those that were included.

The same problem occurs in the study by Subramanian and Kumar (2021), who analyzed only the bivariate relationship between vaccination rates and the number of Covid cases, but omitted large structural differences between countries in the factors that significantly influence the spread of COVID and the public health consequences. Let us illustrate what bias this might have in terms of the impact of vaccination on the spread of Covid infection. To show this, we estimate two models. First, we estimate a simple bivariate model by regressing the logarithm of the 7-day moving average number of new cases per 1 million population on the logarithm of the three-week lagged vaccination rate. In the next step, we estimate a full model that also includes other structural factors (such as the age structure of the population, population density, the prevalence of other diseases that increase the risk of developing severe COVID symptoms (such as diabetes or cardiovascular disease, etc., the availability of quality health care), the current state of the pandemic, and an indicator of countries’ response to epidemics (the stringency index). Figure 2 shows the data and linear regression lines for the bivariate relationship between vaccination rates and the number of new Covid infections. In the simplest model when all structural differences and policy responses of countries are omitted, the model yields a positive and significant relationship (see red regression line trending upwards), i.e. it shows that high vaccination rates lead to a higher number of Covid infections. However, when structural differences and country policy responses are included in the model, the relationship between vaccination rates and the number of new Covid infections becomes negative and significant (see orange regression line trending downwards).

**Figure 2:**
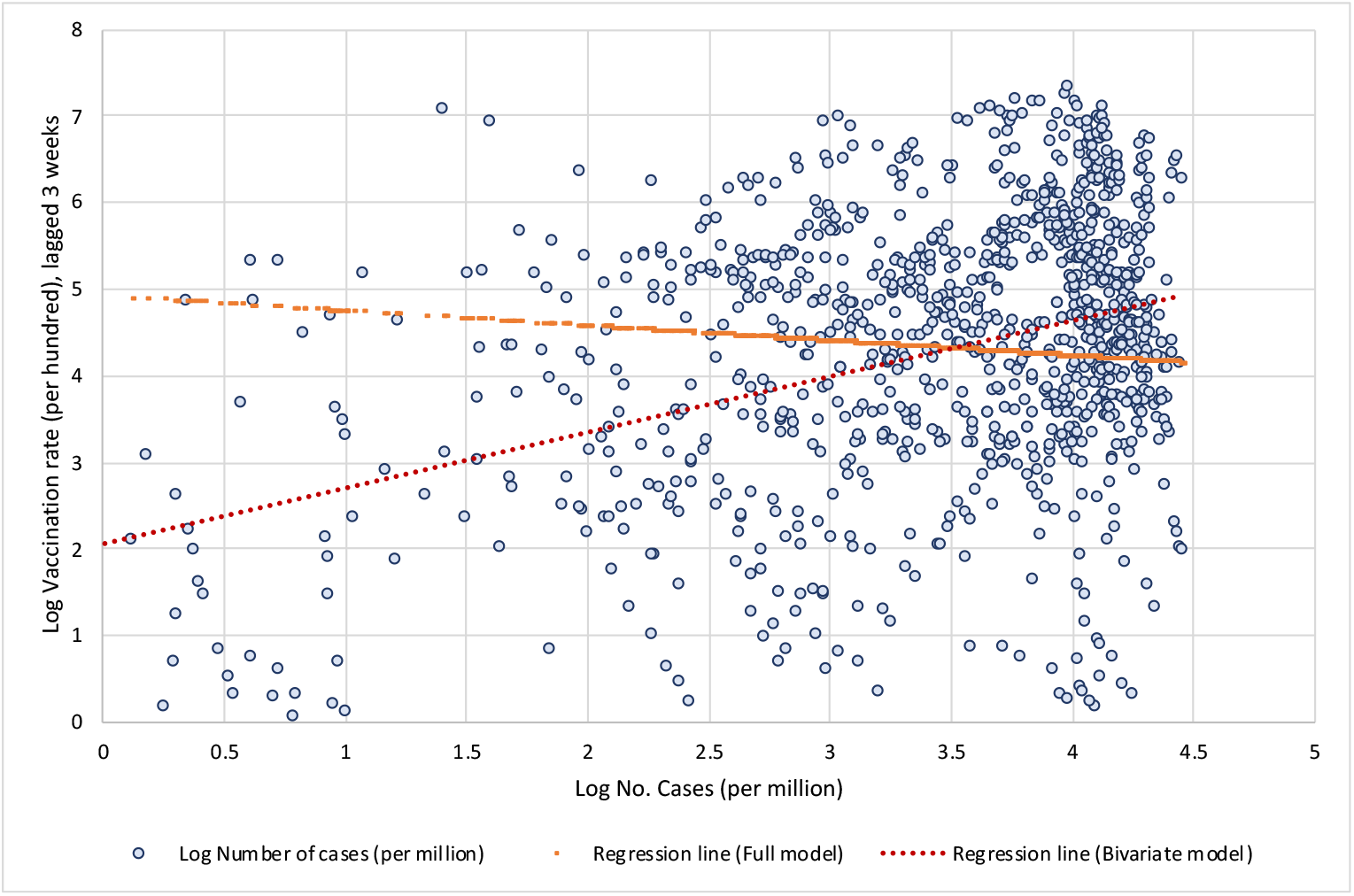
Log Vaccination rate and log Number of cases per million *Notes:* Based on results presented in Table A1 in the Appendix. Data by weeks, period August 1 to November 14, 2021, for 110 countries. Data is specified in natural logarithms; data on vaccination is lagged by 3 weeks. For the Full model the constant term was scaled up. *Source*: Our World in Data; own calculations.

Both issues illustrated above, show that investigating the impact of vaccination on the transmission of COVID in a cross-country setting requires careful analysis. First of all, there are large structural differences between countries in underlying factors that significantly influence the spread of COVID and the public health consequences. In addition, there is wide variation in the response of countries to the pandemic in terms of policies to reduce transmission of COVID, effectiveness in vaccination roll out, etc. There are also large differences between countries in terms of the state of epidemics. One should also consider the timing of vaccination, as immunity takes time to build up, while its effectiveness also gradually wanes after the vaccination.

In this large cross-country study, we take into account the above factors in examining the impact of vaccination on the transmission of COVID and its public health consequences. To analyze the impacts of vaccination, we estimate two comprehensive models – one for the impact on the spread of COVID infections (in terms of number of new confirmed cases) and another one for the impact on severe COVID disease progression (in terms of number of new hospitalizations, admissions to intensive care and deaths). We take into account both the static cross-section and the dynamic aspects of the COVID pandemic. Our empirical models are designed to capture the differences across countries regarding the state of the pandemic (by including the initial level of infections per 1 million) and its dynamics (by including the most recent weekly reproduction rate). In addition to differences in vaccination rates and their dynamics, our model also includes country-specific factors that account for differences between countries that determine countries’ specific vulnerabilities or strengths to perform in dealing with the pandemic.

To examine the effectiveness of vaccination, we use the data for the latest wave of COVID outbreak (the Delta variant) in most countries (the period from August 1, 2021, onwards), while vaccines were also widely available. We use data for 110 countries (from Our World in Data) on the number of cases, deaths, hospitalized and intensive care patients, the percentage of people fully vaccinated, the reproduction rate, and the policy stringency index – all with daily frequency. The COVID-related daily frequency data (per million people) are then transformed into weekly averages using 7-day moving averages. As a point of reference, we use the data for the last day of the week capturing the average effects of the current week. We combine these data with a number of country-specific indicators available at the annual level, such as the percentage of people aged 70+, life expectancy, population density, GDP per capita, number of hospital beds per thousand, cumulative excess mortality per million, diabetes prevalence rate, and cardiovascular death rates.

Our data are structured as a panel with a cross-sectional (country) and time (week) dimension. Both models are estimated using a pooled ordinary least squares (OLS) estimator. We first estimate our baseline models and then proceed with two alternative specifications to examine vaccination effectiveness from a different perspective.

The results can be summarized as follows. First, we show that, after controlling for the number of previous infections, the reproduction rate, the stringency of government protection measures and structural country-specific factors, the full vaccination rate is consistently and significantly negatively correlated with the number of new COVID cases. The regression coefficients indicate that, on average, at a lag of two weeks after the second dose, a 10 percent increase in vaccination rate is associated with a 1.3 percent decrease in new COVID cases. Extending the time elapsed after full vaccination results in a greater decrease in the number of new infections. The overall vaccination effectiveness, however, varies widely from region to region.

Second, our estimations show that magnitude of vaccination contributes significantly to reducing hospitalizations, intensive care (ICU) admissions and deaths with COVID. On average, after controlling for the number of previous infections and structural country-specific factors, a 10 percent increase in the rate of vaccination leads to a reduction of about 5 percent in the number of new hospitalizations, 12 percent decrease in the number of new intensive care patients and 2 percent reduction in the number of new deaths with COVID. Again, there are large differences in vaccination effectiveness between regions. The effects increase the longer the period after full vaccination.

Third, the estimates confirm that the moderating effect of vaccines on the number of cases and deaths occurs when the overall full vaccination coverage is sufficiently high.

Finally, by comparing the data for the same period between 2020 and 2021, we also check the viability of vaccination as a substitute for lockdowns or other, less stringent government protection measures. To do so, we test whether the availability of vaccines has helped countries curb infections and cases of severe disease progression compared to the same period in the previous year when vaccines were not available. More specifically, we check whether the dynamics of the COVID pandemic in the fall of 2021 as compared to the same period in 2020 is moderated in countries with high vaccination rate compared to countries with lower vaccination coverage. Our results suggest that vaccination does not appear to be an effective substitute for more stringent government safety measures to contain the spread of COVID infections until a certain vaccination coverage threshold has been achieved. The spread of infections is shown to be significantly reduced compared to the same period in 2020 only in countries with high vaccination coverage (more than 70 percent).

On the other hand, vaccination is reasonably effective in limiting more severe course of the disease in symptomatic patients already at moderate vaccination rates, as it greatly reduces the number of people requiring hospitalization and intensive care, as well as the number of deaths. Here, moderate vaccination coverage (between 40 and 70 percent) seems to be already a fairly effective tool and can serve in part as a substitute for more stringent government protective measures to reduce the pressure on the health care system.

The outline of the study is as follows. Next section discusses study design, data and empirical methodology. Section 3 present baseline results and results with alternative model specifications. The last section concludes.

## 2. Study design, data and methodology

### 2.1. Study design

The primary goal of COVID vaccination is to protect people against the severe progression of the COVID disease and its public health consequences, i.e. to reduce the number of hospitalizations and the number of people in need of intensive care, and ultimately reduce the number of deaths due to COVID disease. However, the extent to which vaccines reduce transmission of COVID is key to containing the pandemic, which depends on the capacity of vaccines to protect against the spread of the virus. For this reason, we begin our research by first analyzing the impact of vaccination on the spread of new COVID cases and then proceed to study the effectiveness of vaccination in reducing hospitalizations, intensive care unit (ICU) admissions and deaths with COVID.

Studying the impact of vaccination on transmission of COVID in a cross-country setting requires careful consideration. Countries vary considerably in terms of the state of the COVID pandemic, vaccination uptake, policies to reduce transmission COVID, age structure of population, the extent of diseases that increase the risk of developing severe COVID symptoms (such as the prevalence rate of diabetes and the prevalence and severity of cardiovascular disease, etc.), the availability of quality health care, etc. All these and a number of other factors affect the dynamics of COVID epidemics as well as its severity. It is therefore of utmost importance to control for all relevant factors, but also to capture their impact on the dynamics of COVID epidemics.

The analysis should also take into account the timing of vaccination. First of all, immunity takes a while to build up. According to population-based evidence,^9^ there is a lag of at least two weeks after the second dose before the vaccine becomes fully effective in protecting against symptomatic disease. One should also allow for additional lags to account for the lagged effects of vaccination on protection against more severe development of COVID disease, such as an additional week for protection against hospitalization and an additional two to three weeks for protection against mortality. For this reason, in our study, we control for the lagged effects of full vaccination (defined by vaccination with two doses) by allowing for a lag of two to four weeks after full vaccination in estimating the model of the effects of vaccination on the number of new COVID cases and a lag of three to five weeks after full vaccination in estimating the model of the effects of vaccination on hospitalizations, intensive care admissions, and mortality.

There is some evidence of the waning of protection against infection and symptomatic disease over time. Observed protection for fully inoculated is shown to weaken after 15+ weeks since the second dose (to about 45 to 50 percent with AstraZeneca and 70 to 75% with Pfizer-BioNTech and Moderna vaccine). Protection against severe disease and mortality, however, is shown to remain high for at least 5 months after the second dose in most groups (at about 80 to 90 percent with AstraZeneca and 90 to 95% with Pfizer-BioNTech and Moderna vaccine).^10^ These waning effects of vaccination may not affect our results, as countries have built up substantial proportions of the fully vaccinated by early summer, while our data cover the period between August 1 and November 14, 2021.

The design of this study is as follows. To analyze the impact of vaccination on COVID, a comprehensive model must be estimated. As it was demonstrated above, not only are there differences in infection levels between high and low vaccination countries, but there are also differences in the rates of increase in the number of COVID cases. Both aspects, the static cross-section and the dynamic one, need to be considered.

Our empirical model is designed to capture the differences across countries regarding the state of the pandemic (by including the initial level of infections per 1 million) and its dynamics (measured by the past week reproduction rate). In addition to differences in vaccination rates and their dynamics, our model also includes country-specific factors that account for differences between countries that determine countries’ pandemic performance.

To study the effectiveness of vaccination, we chose the period from August 1, 2021, onwards (the most recent data available was until November 14, 2021), as this is the period when the last wave (the Delta variant) of COVID infections reappeared in most countries and when countries had vaccines available. We collect data on the number of cases, deaths, hospitalized and intensive care patients, the percentage of people fully vaccinated, the reproduction rate, and the policy stringency index – all with daily frequency. The last day of the week (Sunday) was used as a reference day in the period capturing the average effect of the previous week. We combine these data with a number of country-specific indicators available at the annual level, such as the percentage of people aged 70+, life expectancy, population density, GDP per capita, number of hospital beds per thousand, cumulative excess mortality per million, diabetes prevalence rate, and cardiovascular death rates.^11^

We first estimate our baseline models to test whether the dynamics of COVID epidemics are mitigated in countries with high vaccination coverage. In addition, we also estimate two alternative specifications to examine vaccination effectiveness from a different perspective. The first test examines whether there is a threshold for vaccination coverage above which vaccines are more effective in moderating the spread of infections and deaths. In the second test, we examine whether the availability of vaccines has helped countries curb the spread of infections and mitigate severe disease progression compared to the same period in the previous year when vaccines were not available. More specifically, we test whether the dynamics of COVID epidemics in the fall of 2021 is moderated in countries with high vaccination coverage compared to countries with low vaccination coverage as compared to the same week in 2020.

### 2.2. Methodology

Unlike observational longitudinal cohort studies,^12^ we apply more rigorous statistical methods using the country-level data to study the effectiveness of vaccination levels across countries on the dynamics of the levels of new COVID cases, new hospitalizations, new ICU patients and new deaths with COVID. To this end, we design two empirical models to be estimated using the detailed country-level data as specified in the next Section.

The first model estimates the impact of vaccination levels on the spread of infections across countries after controlling for other factors. We estimate an exhaustive model that relates the number of new COVID cases to the initial number of COVID cases, the lagged average weekly reproduction rate, full vaccination rate, lagged by 2 to 4 weeks, and a number of country-specific indicators, such as a percentage of people aged 70+, life expectancy, population density, GDP per capita, stringency index, number of hospital beds per thousand, diabetes prevalence rate and cardiovascular death rate.

Obviously, population density is an indicator of the number of potential contacts between people in different countries, while the percentage of people aged 70, life expectancy, diabetes prevalence, and cardiovascular mortality rates account for the susceptibility of the population for more severe consequences of COVID infections. The number of hospital beds per thousand is an indicator of the availability of quality health care, while GDP per capita controls, among other things, for the ability of countries to effectively implement the vaccination campaign. Finally, the stringency index measures the stringency of government action to curb the spread of new infections.

The following model is estimated:

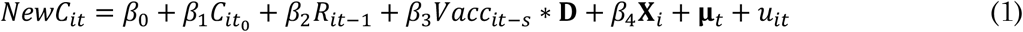

where *NewC*_*it*_ is a dependent variable defined as an end-of-the-week 7-day moving average of the number of new COVID cases per million people in country *i* and week *t* (*t* = 1, …, 16). Our main explanatory variable of interest is the vaccination rate, *Vacc*_*it*−*s*_, which is defined as end-of-the-week 7-day moving average full vaccination rate for each country lagged by the period *s* (*s* = 2, 3, 4). With two to four lags (weeks after full vaccination) we control for lagged effects of differences in the levels of vaccination between countries on mitigating the spread of infections. The vaccination variable is interacted with vector **D** capturing regional dummy variables, with regions defined by continents (Europe, Asia, North America, South America, Oceania). The control group are African countries. By using these interaction terms, we check for any significant differences in the effectiveness of vaccination roll out across continents.

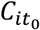 refers to the initial number of new COVID cases defined as 7-day average of the per-million number of COVID cases at the end of the first week in our data sample (*t* = 1). This variable controls for differences in the initial levels of infection between countries. *R*_*it*_ is the end-of-the-week reproduction rate lagged by 1 week, which captures differences in the weekly dynamics of infections between countries. Vector **X**_*i*_ captures all included country-specific indicators, such as a percentage of people aged 70+, life expectancy, population density, GDP per capita, stringency index, number of hospital beds per thousand, diabetes prevalence rate and cardiovascular death rate. Finally, **μ**_*t*_ refers to time (week) fixed effects, while *u*_*it*_ is the remaining i.i.d. error term.

The second model estimates the effectiveness of vaccination levels across countries in protecting against severe COVID disease and its consequences. In this model we relate the number of either new hospitalizations, new ICU patients or new deaths with COVID to the lagged number of COVID cases, full vaccination rate, lagged by 3 to 5 weeks, and several country-specific indicators, such as a percentage of people aged 70+, life expectancy, population density, GDP per capita, stringency index, number of hospital beds per thousand, diabetes prevalence rate and cardiovascular death rate. The following model is estimated:

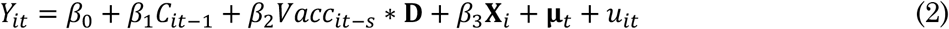

where *Y*_*it*_ is a dependent variable defined interchangeably either in terms of new hospitalizations, new ICU patients or new deaths with COVID. It is defined as end-of-the-week 7-day moving average of the per-million number of new hospitalizations, new ICU patients or new deaths with COVID in country *i* and week *t* (*t* = 1, …, 16). Our main explanatory variable, the vaccination rate *Vacc*_*it*−*s*_, is defined as end-of-the-week 7-day moving average full vaccination rate for each country lagged by the period *s* (*s* = 3, 4, 5). With three to five-week lags (weeks after full vaccination) we control for lagged effects of differences in the levels of vaccination between countries on protection against severe COVID disease. Similarly to the previous model, we interact the vaccination variable with a vector **D** capturing regional dummy variables. *C*_*it*−1_ refers to the lagged number of new COVID cases defined as 7-day moving average of the per-million number of COVID cases at the end of the previous week. This variable controls for differences in the levels of infections between countries. Again, vector **X**_*i*_ captures all included country-specific indicators, such as a percentage of people aged 70+, life expectancy, population density, GDP per capita, stringency index, number of hospital beds per thousand, diabetes prevalence rate and cardiovascular death rate. **μ**_*t*_ refers to time (week) fixed effects, while *u*_*it*_ is the remaining i.i.d. error term.

In both models, all variables are defined in logarithms, which allows all estimated coefficients to be interpreted as elasticities.

Both models are estimated using pooled ordinary least squares (OLS) estimator. Our data are structured as panel data with a cross-sectional (country) and time (week) dimension, which violates the assumption of independence of all observations and leads to the possibility that the error term is correlated with individual regressors in the model. In principle, this requires estimation using the panel data estimator, either with random effects (RE) or with fixed effects (FE). In this case, neither estimator is appropriate. Using the FE estimator would be preferable, but since we include country-specific variables at annual frequency, using the FE estimator would result in dropping all country-specific variables from the estimates. On the other hand, the RE estimator assumes there are unique, time-constant attributes of countries that are not correlated with the individual regressors, which is clearly not the case here. To address this issue, we therefore assume that all country fixed effects are captured by the time-invariant country-specific variables included in the model. Moreover, as the theory suggests, pooled OLS including time fixed effects can be used to derive unbiased and consistent parameter estimates.

We first estimate our baseline models (1) and (2) and then provide several robustness checks to our baseline estimations.

### 2.3. Data collection

We use COVID-19 data and other country-specific data collected by the Our World in Data.^13^ The dataset includes daily data on COVID (COVID-19) number of cases, deaths, hospitalized and ICU patients, number of people fully vaccinated, reproduction rate and stringency index.^14^ This data is combined with a number of country-specific indicators reported at the annual level, such as total population, percentage of people aged 70+, life expectancy, population density, GDP per capita, number of hospital beds per thousand, cumulative excess mortality per million, diabetes prevalence rate and cardiovascular death rate.

The COVID-related daily data was selected for the period August 1 to November 14, 2021,^15^ and for the same period in 2020. This period in 2021 was deliberately chosen to study the effectiveness of vaccination during the period when the last wave of COVID infections hit most countries, while the same period in 2020 is used as control data for the period when COVID vaccines were not available. First, we calculated all COVID-related daily data per 1 million population and then calculated the 7-day moving average for each variable of number of cases, deaths, hospitalizations, and intensive care patients. Next, we kept only the data for the last day in the week (Sunday) in the period from August 1 to November 14, 2021 (i.e., the 7-day moving average data for the past week), effectively giving us up to 16 data points for most countries.

For the vaccination rate, COVID reproduction rate and stringency index, we use the data for the last day in the week in the same period. All other data in our empirical model are annual and therefore there is no variation during the study period.

Our final dataset consists of 110 countries for which data were available for all variables in our empirical model for at least 6 consecutive weeks. In its simplest form, our model is estimated at 1,324 observations. However, the inclusion of up to 5 lags for some of the variables reduces the effective data sample to between 900 and 1,100 observations. Summary statistics is presented in Table A1 in the Appendix.

## 3. Results

We first present our baseline results for model (1) and model (1) and then proceed by testing two alternative model specifications.

### 3.1. Baseline results

#### 3.1.1. Impact of vaccination on number of COVID cases

Our baseline results of estimating model (1) regarding the impact of vaccination on the number of new COVID cases are presented in Table 1. The fit of the model is quite good, as the explanatory variables can explain almost 70 percent of the cross-country and cross-time variation in the new COVID cases. The results show that the full vaccination rate is consistently and significantly negatively correlated with the number of new COVID cases (in the period from August 1 to November 14, 2021). The regression coefficients show that at a lag of 2 weeks after the second dose, a 10 percent increase in vaccination rate is associated with a 1.3 percent decrease in new COVID cases. Increasing the number of weeks after full vaccination results in a greater decrease in the number of new infections - up to a 1.7 percent decrease with a lag of 3 weeks after the second dose.

**Table 1:** Impact of vaccination rate on number of new Covid cases

|  | (1)<br>Lag 2<br>weeks | (2)<br>Lag 3<br>weeks | (3)<br>Lag 4<br>weeks |
| --- | --- | --- | --- |
| Initial No. cases per mill. | 0.609<br>[18.40]*** | 0.577<br>[15.87]*** | 0.571<br>[16.27]*** |
| Reproduction rate_1 | 6.675<br>[16.66]*** | 6.755<br>[15.04]*** | 7.160<br>[16.16]*** |
| Full Vacc rate | -0.130<br>[-2.21]** | -0.172<br>[-2.52]** | -0.157<br>[-2.36]** |
| Full Vacc rate x EU | -0.015<br>[-0.48] | -0.011<br>[-0.33] | -0.030<br>[-0.83] |
| Full Vacc rate x Asia | 0.064<br>[1.58] | 0.082<br>[1.88]* | 0.070<br>[1.54] |
| Full Vacc rate x NorthAm | 0.094<br>[1.95]* | 0.109<br>[2.14]** | 0.085<br>[1.62] |
| Full Vacc rate x SouthAm | -0.067<br>[-1.57] | -0.079<br>[-1.76]* | -0.116<br>[-2.50]** |
| Full Vacc rate x Oceania | -0.024<br>[-0.28] | -0.034<br>[-0.40] | -0.050<br>[-0.57] |
| Aged 70+ (percent) | 0.570<br>[6.36]*** | 0.631<br>[6.40]*** | 0.666<br>[6.57]*** |
| Life expectancy | 0.143<br>[0.14] | -0.355<br>[-0.27] | -0.308<br>[-0.24] |
| Population density | -0.025<br>[-0.83] | -0.044<br>[-1.34] | -0.054<br>[-1.51] |
| GDP per capita | 0.369<br>[4.00]*** | 0.408<br>[4.03]*** | 0.426<br>[4.12]*** |
| Stringency index | 0.331<br>[2.83]*** | 0.281<br>[2.37]** | 0.356<br>[3.18]*** |
| No. hospital beds | 0.004<br>[0.06] | 0.024<br>[0.36] | 0.045<br>[0.65] |
| Cardiovasc. death rate | 0.481<br>[4.76]*** | 0.452<br>[4.13]*** | 0.489<br>[4.39]*** |
| Diabetes prevalence | 0.157<br>[1.40] | 0.178<br>[1.47] | 0.234<br>[1.87]* |
| Constant | -11.819<br>[-2.63]*** | -9.585<br>[-1.72]* | -10.843<br>[-1.97]** |
| Observations | 1,088 | 1,002 | 914 |
| R-squared | 0.667 | 0.668 | 0.684 |

*Notes:* Dependent variable is defined as logarithm of an end-of-the-week 7-day moving average of the per-million number of new COVID cases. The lag of full vaccination rate is indicated in the head of the column. All variables are specified in natural logarithms. The model is estimated by Pooled OLS and includes time fixed effects.
Robust t-statistics in brackets, \*\*\* p<0.01, \*\* p<0.05, \* p<0.1.

The overall vaccination effectiveness varies widely from region to region. While countries in the European Union (EU) and Oceania are not significantly different from the average, the overall effectiveness of the vaccination in containing the spread of infections in countries of Asia and North America is only about half the average.^16^ In South American countries, on the other hand, higher vaccination rates lead to about 50 percent greater impact on reducing the number of new COVID infections.

Other variables in the model yield expected results. As predicted, new COVID infections are significantly positively correlated with the initial number of infections and the reproduction factor, confirming the obvious impact of the pandemic and its dynamics on future infections.

Among structural factors, the proportion of the population aged 70+ has a significant positive impact on the spread of infections. Life expectancy is not significantly correlated with the number of new covid cases, most likely because its impact was in part absorbed by the proportion of the population aged 70+. Population density also does not seem to be associated with the spread of infections. On the other hand, GDP per capita is positively correlated with the number of new cases, confirming that COVID -19 is a disease of rich countries. The quality of health care, expressed by the number of hospital beds per capita, is also not correlated with the magnitude of new COVID cases, as its effect was likely picked-up by GDP per capita, which controls for the level of development. The positive correlation between the stringency index and the number of new cases indicates that government policies correspondingly react to the surge in new infections to contain the spread of the virus, but are unable to contain it quickly. As expected, the increase in new COVID cases is significantly positively associated with cardiovascular death rate, while diabetes prevalence is correlated only in some specifications.

We also estimate the impact of vaccination separately for European Union (EU) countries, which have adopted a common strategy to secure supplies and facilitate distribution of vaccines. This led to effective vaccine distribution, with more than 75 percent of the population aged 18 and older fully vaccinated by mid-November 2021. However, not all EU countries have been equally successful in vaccinating their populations. An interesting pattern can be seen along the former “Iron Curtain”: Western EU Member States were more successful in their efforts to fully vaccinate their populations, while Eastern EU Member States did not keep up. This is reflected in two extremes: on the one hand, Ireland has fully vaccinated more than 92 percent of its over-18 population, while in Bulgaria only 29 percent of adults were fully vaccinated as of mid-November 2021.

In this sense, it is interesting to investigate how these differences in vaccination roll out impacted at the dynamics of COVID outbreaks in different countries in autumn 2021. We estimate a model (1) where we add an interaction term for the full vaccination rate and the countries exceeding the average vaccination rate in the EU. The results show a very strong negative response of new COVID cases to the level of vaccination. On average, after controlling for the state of the pandemic and structural country-specific factors, a 1 percent increase in vaccination rate is associated with a 1.07 to 1.35 percent reduction in the number of new cases in the 2 to 4 weeks following full vaccination. In addition, in EU countries with above-average vaccination rates, the increase in new COVID cases is smaller by an additional 0.13 to 0.15 percent for each percent of above-average vaccination rate (see Figure 3). This confirms the importance of vaccination efforts by EU countries to curb the momentum of COVID infections.

**Figure 3:**
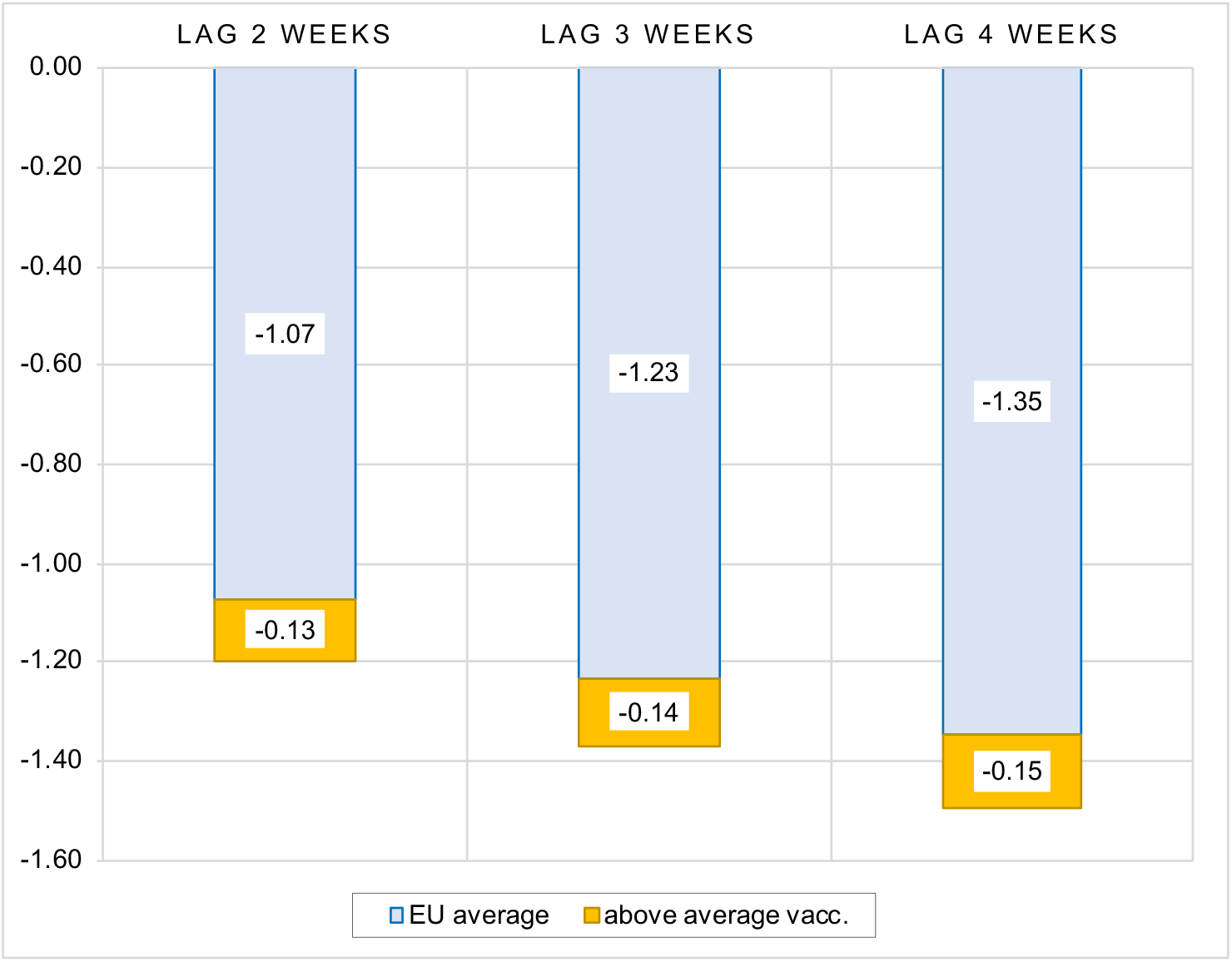
Impact of vaccination rate on number of new Covid cases, sample of EU countries *Notes:* Figure contains the estimated elasticities of the per-million number of new COVID cases to the vaccination rate obtained by estimating the model (1). See Table A3 in the Appendix for the full results.

**Figure 3:**
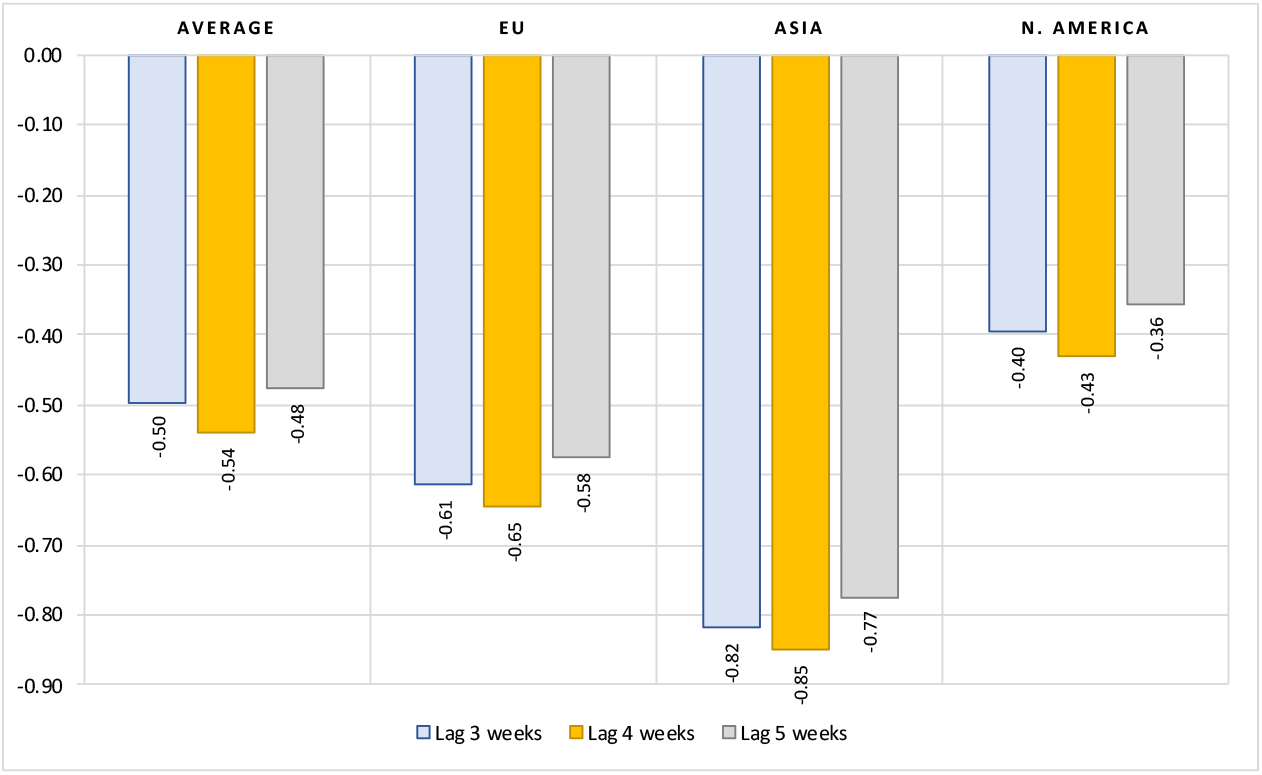
Impact of vaccination rate on number of new hospitalized patients, limited data sample* *Notes:* Figure contains the estimated elasticities of the per-million number of new hospitalized patients to vaccination rate obtained by estimating the model (2). (*) Data available only for most of European countries, United States, Canada and Israel. See Table A4 in the Appendix for the full results.

#### 3.1.2. Impact of vaccination on number of hospitalizated and ICU patients

In this section, we draw inferences about how effective the differences in COVID vaccination across countries are in protecting against severe COVID disease, i.e., in curbing the number of hospitalizations and the number of people requiring intensive care (ICU) treatment. We estimate model (2) using data for all countries with available data. Note that data for hospitalizations and ICU admissions are available only for advanced countries, i.e. for most, but not all, European countries, United States, Canada and Israel, which means that our data sample shrinks to only one-third of the sample used in the previous section (from 1,000 to about 300 weekly observations). Therefore, the results are not directly comparable to those obtained for the impact of vaccination on the number of new cases, as the data samples differ in both number of observations and geographic coverage. For the sake of brevity, we present only the figures with the main variables of interest, i.e. the elasticity of new hospitalized and ICU patients to vaccination rate and the regional interaction terms, while the full estimation results can be found in the Appendix.

The results in Figure 3 show that the magnitude of vaccination contributes significantly to reducing hospitalizations due to COVID. On average, after controlling for the number of active cases and structural country-specific factors, a 10 percent increase in the rate of vaccination leads to about a 5-percent reduction in the number of new hospitalizations. The impact is greatest three weeks after full vaccination. This effect is stronger by about 20 to 30 percent in the EU (vaccination elasticity of about 0.6 to 0.65) and by about 60 percent in Asian countries (vaccination elasticity of about 0.8). In North American countries, on the other hand, the elasticity of the number of new hospitalized patients to the vaccination rate is much lower - only about 0.4.^17^

Figure 4 presents estimated elasticities for the impact of vaccination rate on number of new ICU patients. The results show that COVID vaccination, with an elasticity of about 1.2, is even more effective in reducing the number of patients requiring intensive care. For example, on average, a 10 percent increase in vaccination rates results in about a 12 percent decrease in the number of new intensive care patients due to COVID. There are no significant differences in this effect in the EU and Asian countries, while this vaccination effect is about 20 percent lower in North America (vaccination elasticity of about 1).

**Figure 4:**
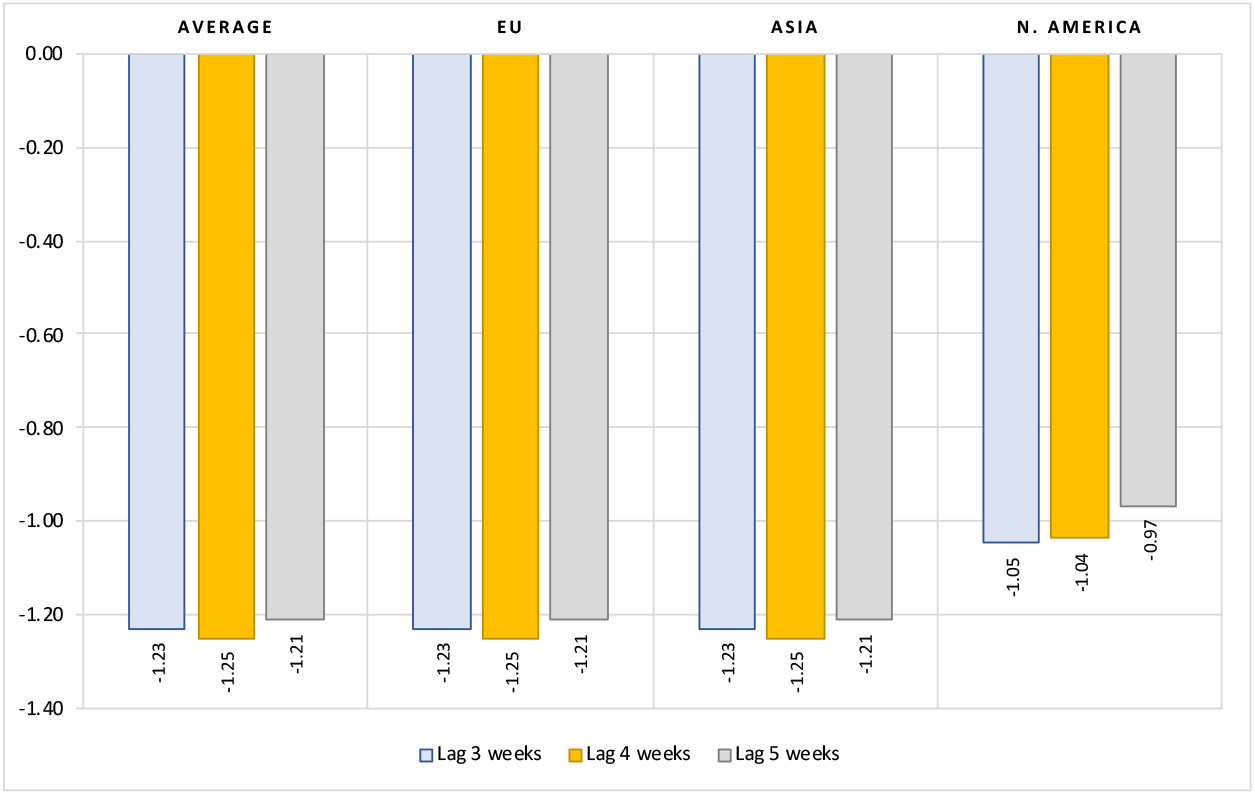
Impact of vaccination rate on number of new ICU patients, limited data sample* *Notes:* Figure contains the estimated elasticities of the per-million number of new ICU patients to vaccination rate obtained by estimating the model (2). (*) Data available only for most of European countries, United States, Canada and Israel. See Table A5 in the Appendix for the full results.

#### 3.1.3. Impact of vaccination on number of new deaths with COVID

Finally, we also examine how effective differences in COVID vaccination rates between countries are in reducing deaths associated with COVID. Again, we only show the elasticities for the main variables of interest, while the full results can be found in the Appendix. Results in Figure 5 confirm that after controlling for the number of previous infections and structural country-specific factors, countries with higher vaccination coverage against COVID perform better in terms of lives saved, although the elasticities are small in magnitude. On average, a 10 percent increase in full vaccination coverage leads to about a 2 percent reduction in the number of new deaths with COVID. The effect increases the longer the period after full vaccination. In Asia and Oceania, these effects are no different from the average effects, while in the EU and North America, the impact of the extent of vaccination on the reduction in the number of deaths is much smaller. In EU countries, a 10 percent increase in vaccination coverage is associated with a 0.8 to 1 percent decrease in new deaths, while in North American countries the response to a 10 percent increase in vaccination coverage is 0.4 to 0.6 percent. In both regions, these effects diminish in the weeks following full vaccination. This likely indicates the gradual waning of the vaccination effect in these two regions, which began early in the vaccination campaigns.

**Figure 5:**
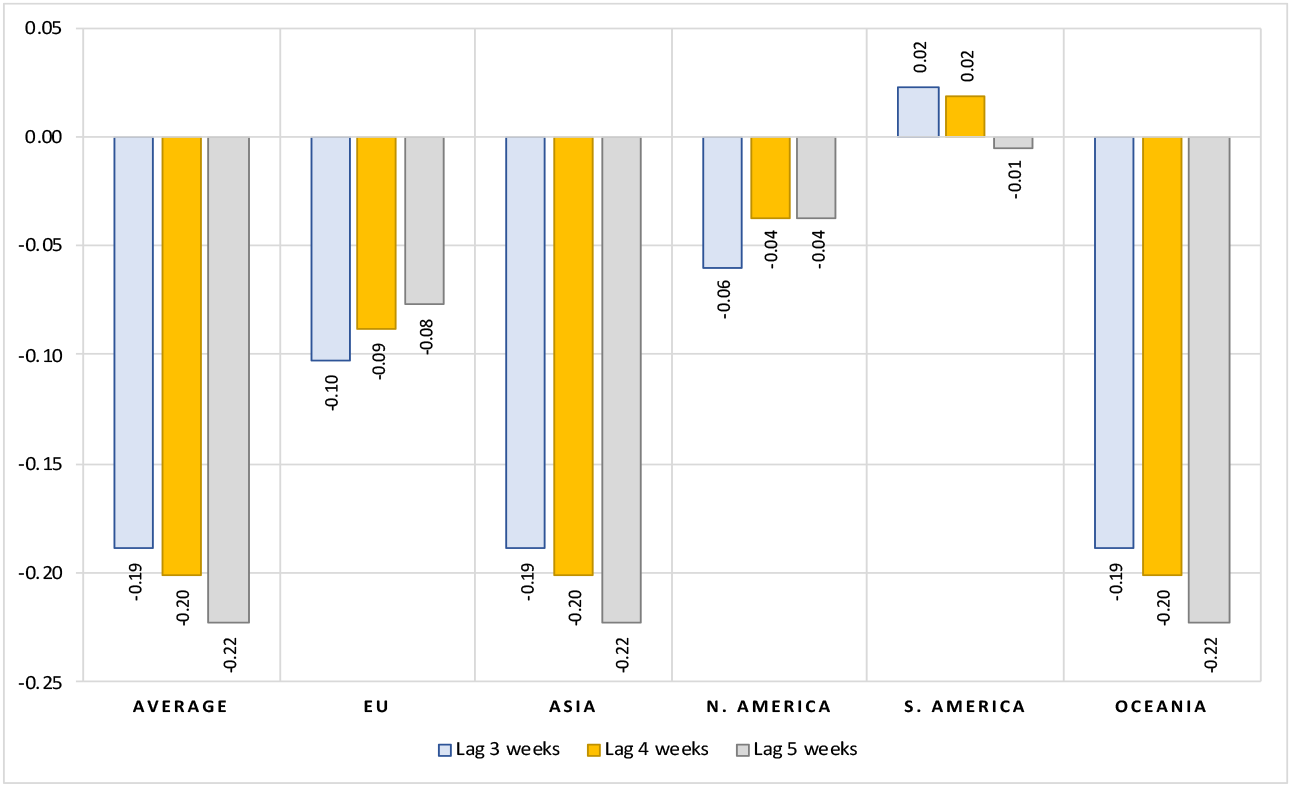
Impact of vaccination rate on number of new deaths, all countries *Notes:* Figure contains the estimated elasticities of the per-million number of new deaths with COVID to vaccination rate obtained by estimating the model (2). See Table A6 in the Appendix for the full results.

### 3.2. Alternative specifications

The results presented so far clearly indicate that vaccination campaigns have been quite effective in containing the spread of infection and reducing the number of people with severe cases of the disease and eventually deaths. In what follows, we present two alternative model specifications to test the effectiveness of vaccines from a different angle. In the first robustness check, we test whether there is some threshold in vaccination coverage at which vaccines become more effective in moderating the spread of infections and deaths. In the second alternative specification, we test how effective vaccination was in curbing the spread of infection and symptomatic disease progression compared to the same period in 2020, when vaccines were not yet available.

#### 3.2.1. Is there a threshold of for efficient vaccination coverage?

The evidence from Israel, the United Kingdom, and other Western countries show that despite the more infectious delta variant, the spread of infection in 2021 was subdued in countries with high vaccination coverage of population as compared with 2020. Later in the fall of 2021, as the recent covid wave swelled and new cases reappeared in many European countries, the number of hospitalized patients and deaths remained relatively low in countries with a larger proportion of population inoculated. This obviously indicates that vaccination is only effective in curbing the incidence of new cases, hospitalizations, and deaths when a sufficient vaccination threshold is achieved.

Below we test whether there are differences in vaccine effectiveness between countries with low, medium and high levels of vaccination. We use a similar approach as in Aizenman et al (2021) by forming three groups of countries according to vaccination coverage. The first group consists of countries with less than 40 percent of the population fully vaccinated. The middle group contains countries with vaccination coverage between 40 and 70 percent, while the high vaccination group contains countries with vaccination coverage of more than 70 percent. Based on this grouping, in our empirical models we interact the vaccination rate variable with these three groups of countries. We re-estimate models (1) and (2) by adding two new interaction terms of vaccination rates with dummy variables for medium and high vaccinated countries. The country group with the low vaccination rates is used as the control group.

The results presented in Figure 6 show that the moderating effect of vaccines on the number of cases and deaths occurs when the overall rate of full vaccination is sufficiently high. While the impact of vaccines is always significantly negative, the vaccination threshold for moderating the spread of new infections is quite high for vaccination coverage rates above 70 percent, while it is less efficient in the intermediate group of countries with vaccination coverage rates between 40 and 70 percent. In terms of deaths, countries with low vaccination coverage rates do not seem to benefit significantly from vaccination. Here, the moderating effect of vaccination already becomes effective at moderate vaccination coverage rates and intensifies with increasing vaccination coverage rates and the length of the period after full vaccination.

**Figure 6:**
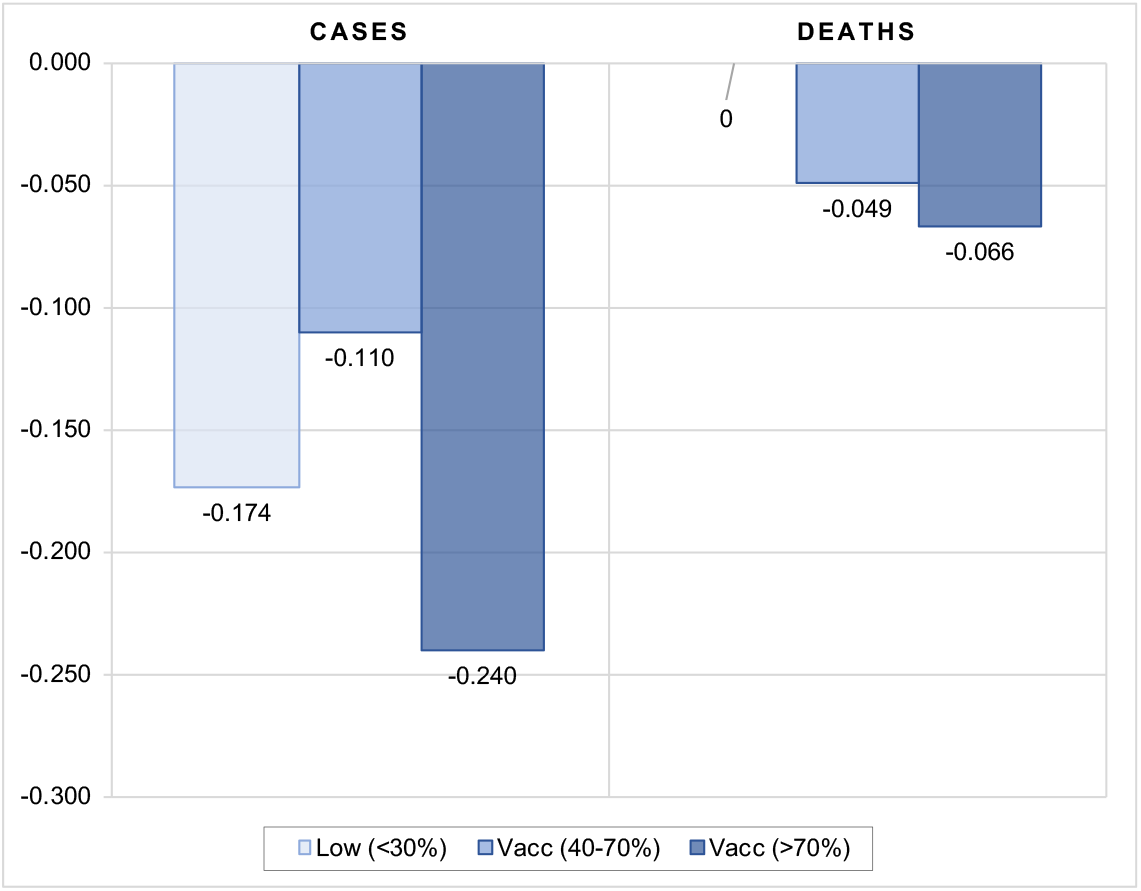
Impact of vaccination threshold rate on number of new cases and deaths, all countries *Notes:* Figure contains the estimated elasticities of the per-million number of new COVID cases and deaths to vaccination rate obtained by estimating the models (1) and (2). The elasticities in the figure are calculated as a sum of vaccination coefficient and the corresponding interaction terms with the vaccination group. The vaccination variable is lagged by 4 weeks. See Table A7 in the Appendix for the full results and different lags structure.

#### 3.2.2. Do vaccines substitute for lockdowns?

In this section, we examine how effective vaccination was in curbing the spread of infection and severe disease progression compared to the same period in 2020, when vaccines were not yet available. In the fall of 2020, most advanced countries resorted to either full or partial lockdown measures, including travel restrictions, school closings, banning public gatherings, closing workplaces, or even imposing a full or nighttime curfew to slow the spread of the virus. Some studies show that lockdown measures had a small but significant effect on reducing the number of cases per million, but a larger effect on mortality (see Violato et al, 2021). In contrast, given the availability of vaccines and despite the outbreak of delta variant in 2021 compared to the same period in 2020, most countries have significantly lifted restrictions on the movement of people (see Figure 7). It is interesting to see to what extent vaccines can serve as a substitute for lockdowns.

**Figure 7:**
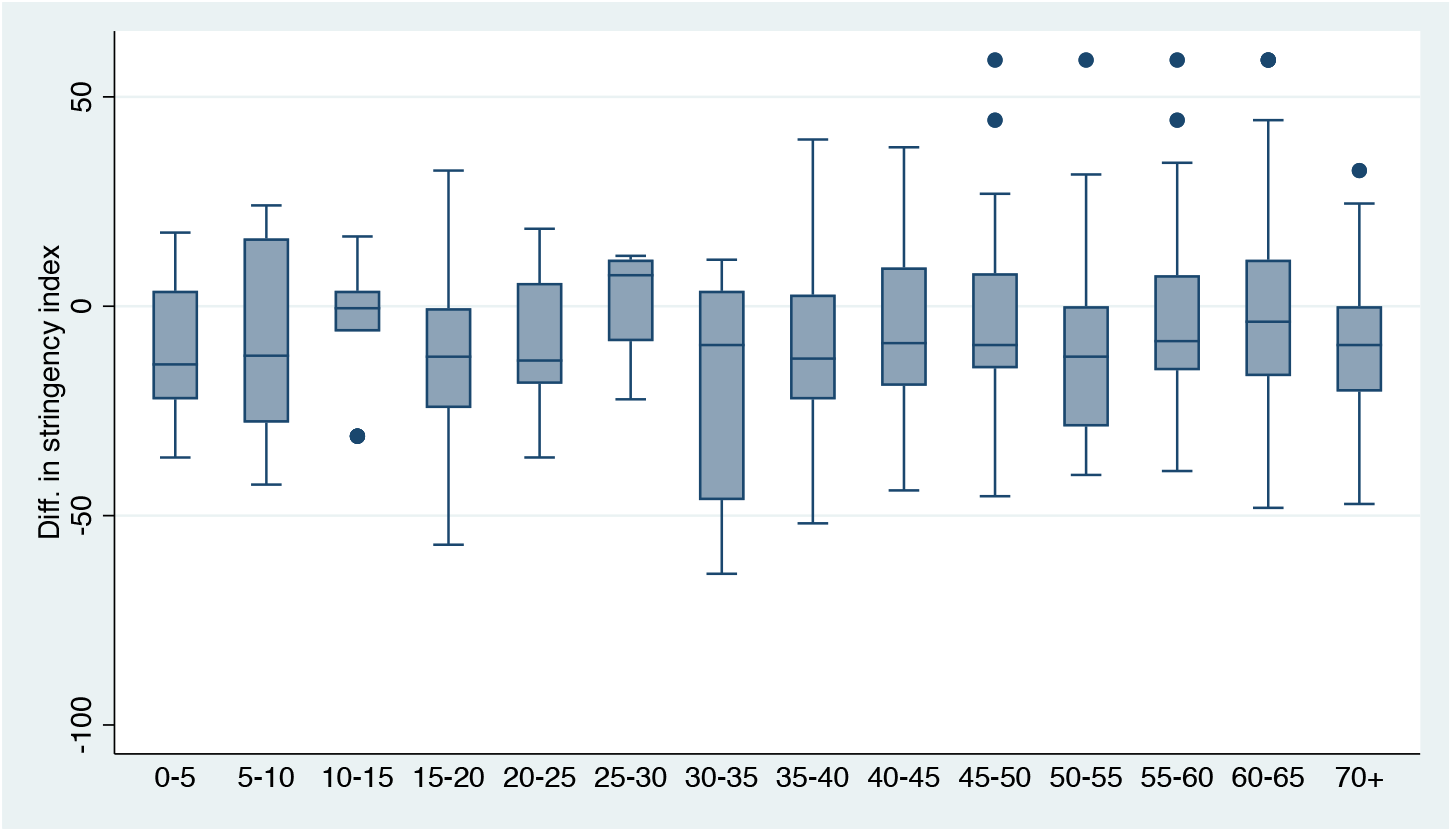
Median, interquartile range and variation in differences in stringency measures between the same period in 2021 and 2020, by vaccination coverage, all countries *Notes:* Vaccination rate bins on the horizontal axis. Underlying data are calculated as differences in 7-day average of stringency index between the same week in 2021 and 2020. Median, interquartile range and variation are shown for the last available 6-week period (October 3 to November 14, 2021 v. October 4 to November 15, 2020) *Source:* Our World in Data; own calculations.

In what follows we formally examine whether in the presence of vaccines this relaxation of government stringency measures in 2021 led to a significant increase in new cases, hospitalizations, ICU admissions and deaths in 2021 compared to the same period in 2020. Specifically, we test whether the dynamics of COVID epidemics in the fall of 2021 were moderated in countries with high vaccination coverage relative to countries with low vaccination coverage compared to the same week in 2020. To do this, we re-estimate our models (1) and (2) using data calculated as differences in the number of confirmed infections, hospitalizations, ICU admissions, and deaths between the same 16 weeks in 2021 and 2020.^18^

The results in Figure 8 indicate that vaccination matters for pandemic dynamics in the absence of more stringent government policies and lockdowns. In countries with low vaccination coverage (below 40 percent), vaccination had no significant impact on the number of new cases compared to 2020, while in countries with medium coverage (between 40 and 70 percent), the number of new cases was actually higher in 2021 compared to the same period in 2020. However, in countries with high vaccination coverage (more than 70 percent), the number of new cases was significantly lower in 2021 compared to the same period in 2020. This indicates that vaccination does not appear to be an effective substitute for more stringent government protection measures to contain the spread of COVID infections as long as vaccination rates are low to moderate.

**Figure 8:**
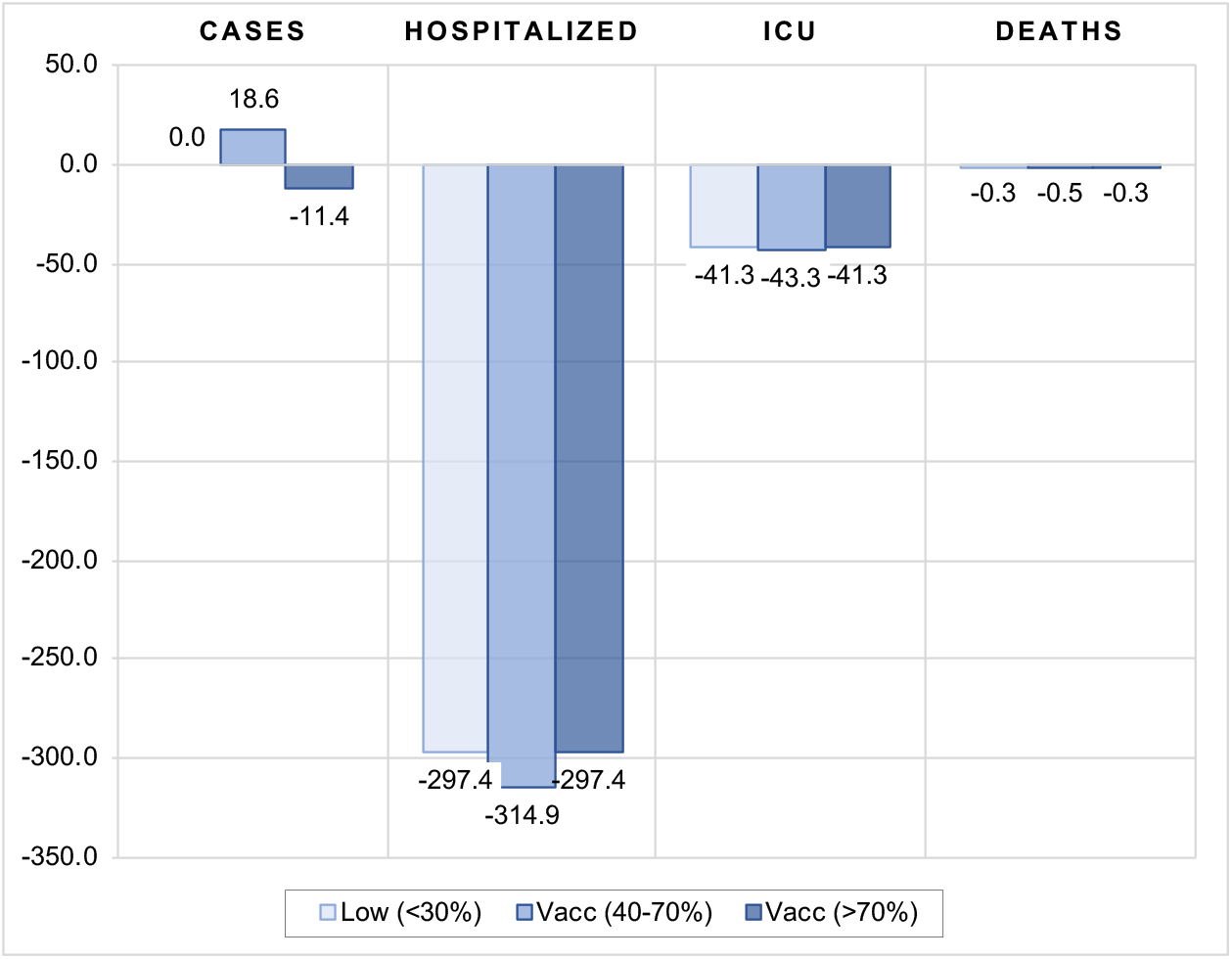
Impact of vaccination rate on number of new cases, hospitalizations, ICU admissions and deaths in 2021 compared to 2020 *Notes:* Figure contains the estimated coefficients of the per-million number of cases, hospitalized and ICU patients, and new deaths with COVID to vaccination rate obtained by estimating the models (1) and (2). The underlying data was calculated as differences in the corresponding variable between the same 16 weeks of August to November in 2021 and 2020.The vaccination variable is lagged by 4 weeks. Note that for new cases and deaths, data were used for the entire country sample, whereas for hospitalized and intensive care patients, data are only available for most European countries, the United States, Canada and Israel. See Tables A8 and A9 in the Appendix for the full results and different lags structure.

On the other hand, our results indicate that vaccination is effective in limiting the more severe course of the disease in symptomatic patients, as it greatly reduces the number of people requiring hospitalization and intensive care, as well as the number of deaths, compared to the same period in 2020. Although the vaccination effect is always significantly negative, the impact is shown to be strongest in countries with moderate vaccination coverage (between 40 and 70 percent). For instance, in countries with low and high vaccination coverage, a 1 percentage point increase in vaccination coverage leads to a decrease in hospital and intensive care patients of 297 and 41 per million population, respectively, and to 0.3 fewer deaths per million population compared to the same period in 2020. In countries with moderate vaccination coverage, these effects are about 5 percent more favorable in terms of number of hospitalized and ICU patients and 40 percent more favorable in terms of lives saved.

These results indicate that moderate-grade vaccination coverage seems to be already an effective tool and can serve in part as a substitute for more stringent government protective measures. Of course, we do not know what the estimated coefficients would be if more countries had reached the high vaccination threshold. We can only speculate that, in this case, higher protection would lead to a smaller number of people with severe disease than would be the case with moderate vaccination coverage.

## 4. Conclusions

This study analyzes the impact of vaccination on the transmission of COVID and its public health consequences. To this purpose, we make use of large cross-country dataset for 110 countries to estimate two comprehensive models of the effects of inoculation – one for the impact on the spread of COVID infections and another one for the impact on the frequency of severe COVID disease progression. In estimating the vaccines effectiveness, the models capture the differences across countries regarding the state of epidemics and its dynamics and country-specific factors. The latter account for differences between countries that determine countries’ specific vulnerabilities or strengths to perform worse or better during epidemics.

To examine the effectiveness of vaccination, we use daily COVID-related data (from Our World in Data) for the period from August 1, 2021, onwards, which captures both the last wave of COVID outbreak (the Delta variant) in most countries and the availability of vaccines.

Our results confirm that vaccines are reasonably effective in both limiting the spread of infections and containing more severe disease progression in symptomatic patients. First, results show that the vaccination rate (of fully inoculated individuals) is consistently negatively correlated with the number of new COVID cases, whereby a 10 percent increase in vaccination rate is associated with a 1.3 to 1.7 percent decrease in new COVID cases. Second, the estimates show that magnitude of vaccination contributes significantly to moderating severe disease progression. On average, a 10 percent increase in the rate of vaccination leads to about a 5 percent reduction in the number of new hospitalizations, 12 percent decrease in the number of new intensive care patients and 2 percent reduction in the number of new deaths with COVID. While the effects increase with time elapsed since full vaccination, the overall vaccination effectiveness is shown also to vary widely from region to region. Third, the estimations show that the moderating effect of vaccines on the number of cases and deaths occurs when the total number of vaccinations is sufficiently high.

And finally, by comparing the data for the same period between 2020 and 2021, we also check how viable is vaccination as a substitute for lockdowns or other less restrictive government measures against the spread of COVID. Our results suggest that vaccination does not appear to be an effective substitute for more restrictive government safety measures to contain the spread of COVID infections until a high vaccination coverage threshold (more than 70 percent) has been achieved. On the other hand, vaccination is shown to be quite effective in limiting the more severe course of the disease in symptomatic patients already at moderate vaccination coverage (between 40 and 70 percent).

In conclusion, our results show that vaccines are quite effective in both limiting the spread of infection and containing a more severe course of disease in symptomatic patients. High vaccination coverage has been shown to be a reasonably effective tool and may serve in part as a substitute for more stringent government protective measures. In this way, it can also help to reduce pressure on the health system and thus benefit the overall public health of society during such severe pandemics.

## Data Availability

All data and code used in the present study are available upon request to the authors

https://www.dropbox.com/t/gUo4G60HXw85EpZC

## Statements

### Author Contributions

All authors contributed to the study conception and design. Material preparation, data collection and analysis were performed by Jože Damijan, Sandra Damijan and Črt Kostevc. The first draft of the manuscript was written by Jože Damijan and all authors commented on previous versions of the manuscript. All authors read and approved the final manuscript.

## Appendix

**Table A1:** Summary statistics

| **Variable** | **Obs.** | **Mean** | **Std. Dev.** | **Min** | **Max** |
| --- | --- | --- | --- | --- | --- |
| Country | 1,824 | 109.2 | 62.6 | 1 | 110 |
| Week | 1,824 | 8.5 | 4.6 | 1 | 16 |
| New cases per mill. | 1,824 | 151.3 | 215.8 | 0 | 1523.1 |
| New deaths per mill. | 1,824 | 1.9 | 3.1 | -0.15 | 25.0 |
| New ICU patients per mill. | 362 | 20.3 | 20.4 | 0 | 106.1 |
| New hospitalized per mill. | 418 | 123.8 | 165.0 | 4.8 | 1213.6 |
| Full Vaccination rate (%) | 1,318 | 41.1 | 23.3 | 0.05 | 87.5 |
| Reproduction rate | 1,710 | 1.0 | 0.3 | 0 | 2.1 |
| Stringency index | 1,712 | 50.9 | 16.3 | 13.9 | 96.3 |
| Population density | 1,824 | 232.7 | 770.7 | 2.0 | 7915.7 |
| Median age | 1,824 | 32.9 | 8.5 | 15.1 | 48.2 |
| Share of population aged 70+ (%) | 1,824 | 6.6 | 4.5 | 0.5 | 18.5 |
| GDP per capita (US \$) | 1,824 | 22524.6 | 20093.5 | 926.0 | 116935.6 |
| Cardiovasc. death rate | 1,824 | 247.4 | 124.7 | 79.4 | 724.4 |
| Diabetes prevalence | 1,824 | 7.6 | 3.5 | 1.8 | 17.7 |
| Number of hospital beds per thousand | 1,824 | 3.2 | 2.5 | 0.3 | 13.1 |
| Life expectancy | 1,824 | 75.0 | 6.2 | 60.2 | 84.6 |

*Source:* Our World in Data; own calculations.

**Table A2:** Impact of vaccination rate on number of new Covid cases

|  | (1)<br>Lag 3 weeks | (2)<br>Lag 3 weeks |
| --- | --- | --- |
| Initial No. cases per mill. |  | 0.577<br>[15.87]*** |
| Reproduction rate_1 |  | 6.755<br>[15.04]*** |
| Full Vacc rate | 0.644<br>[12.74]*** | -0.172<br>[-2.52]** |
| Full Vacc rate x EU |  | -0.011<br>[-0.33] |
| Full Vacc rate x Asia |  | 0.082<br>[1.88]* |
| Full Vacc rate x NorthAm |  | 0.109<br>[2.14]** |
| Full Vacc rate x SouthAm |  | -0.079<br>[-1.76]* |
| Full Vacc rate x Oceania |  | -0.034<br>[-0.40] |
| Aged 70+ (percent) |  | 0.631<br>[6.40]*** |
| Life expectancy |  | -0.355<br>[-0.27] |
| Population density |  | -0.044<br>[-1.34] |
| GDP per capita |  | 0.408<br>[4.03]*** |
| Stringency index |  | 0.281<br>[2.37]** |
| No. hospital beds |  | 0.024<br>[0.36] |
| Cardiovasc. Death rate |  | 0.452<br>[4.13]*** |
| Diabetes prevalence |  | 0.178<br>[1.47] |
| Constant | 2.060<br>[11.32]*** | -9.585<br>[-1.72]* |
| Observations | 1,080 | 1,002 |
| R-squared | 0.153 | 0.668 |

*Notes:* Dependent variable is defined as logarithm of an end-of-the-week 7-day moving average of the per-million number of new COVID cases. Full vaccination rate is lagged by 3 weeks. All variables are specified in natural logarithms. The model is estimated by Pooled OLS and includes time fixed effects.
Robust t-statistics in brackets, \*\*\* p<0.01, \*\* p<0.05, \* p<0.1.

**Table A3:**
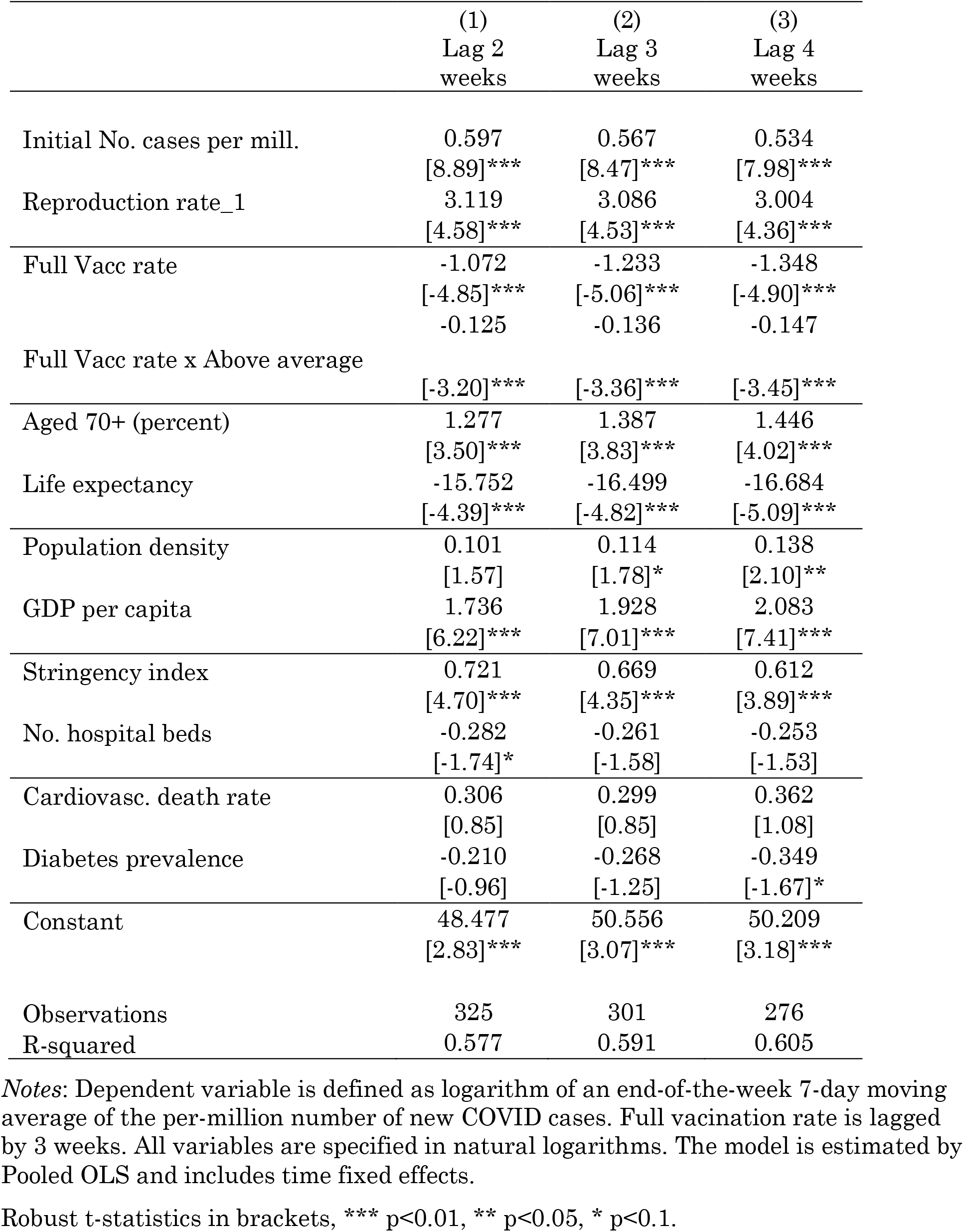
Impact of vaccination rate on number of new Covid cases, sample of EU countries

**Table A4:**
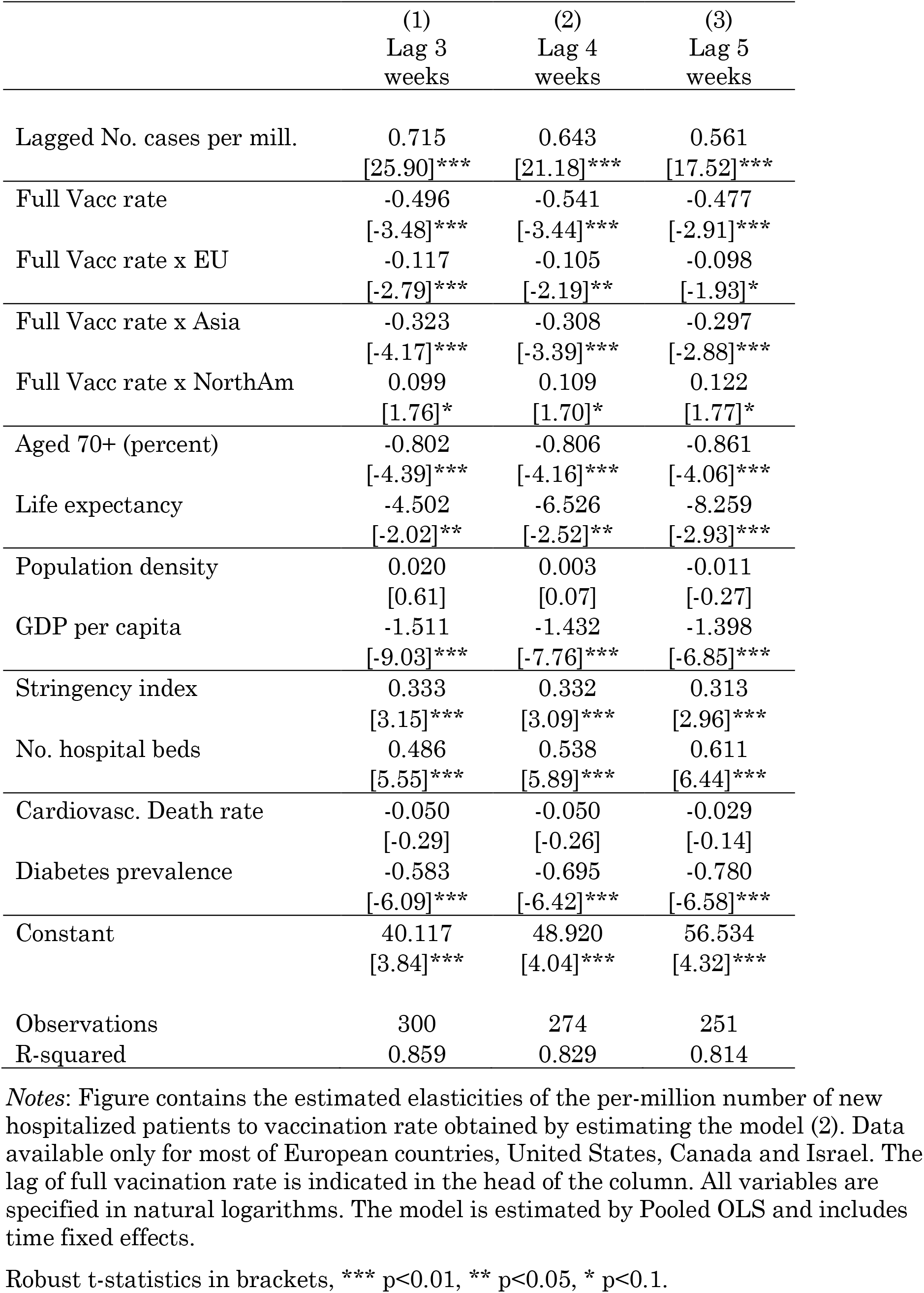
Impact of vaccination rate on number of new hospitalized patients

**Table A5:** Impact of vaccination rate on number of new ICU patients

|  | (1)<br>Lag 3<br>weeks | (2)<br>Lag 4<br>weeks | (3)<br>Lag 5<br>weeks |
| --- | --- | --- | --- |
| Lagged No. cases per mill. | 0.673<br>[13.98]*** | 0.609<br>[10.68]*** | 0.520<br>[8.20]*** |
| Full Vacc rate | -1.229<br>[-6.52]*** | -1.251<br>[-5.26]*** | -1.210<br>[-4.17]*** |
| Full Vacc rate x EU | -0.034<br>[-0.77] | -0.021<br>[-0.39] | -0.009<br>[-0.15] |
| Full Vacc rate x Asia | -0.086<br>[-1.57] | -0.053<br>[-0.79] | -0.030<br>[-0.36] |
| Full Vacc rate x NorthAm | 0.183<br>[3.28]*** | 0.215<br>[3.10]*** | 0.243<br>[2.98]*** |
| Aged 70+ (percent) | -0.178<br>[-0.75] | -0.162<br>[-0.54] | -0.228<br>[-0.61] |
| Life expectancy | -4.183<br>[-1.17] | -5.022<br>[-1.13] | -5.778<br>[-1.04] |
| Population density | -0.004<br>[-0.11] | 0.001<br>[0.02] | 0.001<br>[0.02] |
| GDP per capita | 0.007<br>[0.05] | 0.089<br>[0.44] | 0.096<br>[0.38] |
| Stringency index | 0.520<br>[4.33]*** | 0.506<br>[3.32]*** | 0.471<br>[2.61]*** |
| No. hospital beds | 0.925<br>[10.10]*** | 1.004<br>[9.19]*** | 1.081<br>[8.62]*** |
| Cardiovasc. Death rate | -0.534<br>[-3.05]*** | -0.442<br>[-2.04]** | -0.394<br>[-1.47] |
| Diabetes prevalence | 0.179<br>[1.74]* | 0.097<br>[0.80] | 0.034<br>[0.25] |
| Constant | 22.128<br>[1.47] | 24.897<br>[1.34] | 28.445<br>[1.25] |
| Observations | 262 | 240 | 218 |
| R-squared | 0.840 | 0.790 | 0.752 |

*Notes:* Figure contains the estimated elasticities of the per-million number of new ICU patients to vaccination rate obtained by estimating the model (2). Data available only for most of European countries, United States, Canada and Israel. The lag of full vaccination rate is indicated in the head of the column. All variables are specified in natural logarithms. The model is estimated by Pooled OLS and includes time fixed effects.
Robust t-statistics in brackets, \*\*\* p<0.01, \*\* p<0.05, \* p<0.1.

**Table A6:** Impact of vaccination rate on number of new Covid deaths

|  | (1)<br>Lag 3<br>weeks | (2)<br>Lag 4<br>weeks | (3)<br>Lag 5<br>weeks |
| --- | --- | --- | --- |
| Lagged No. cases per mill. | 0.820<br>[39.09]*** | 0.797<br>[30.46]*** | 0.747<br>[25.23]*** |
| Full Vacc rate | -0.189<br>[-3.90]*** | -0.201<br>[-3.37]*** | -0.223<br>[-3.74]*** |
| Full Vacc rate x EU | 0.086<br>[3.91]*** | 0.113<br>[4.92]*** | 0.146<br>[5.86]*** |
| Full Vacc rate x Asia | -0.002<br>[-0.08] | 0.009<br>[0.35] | 0.016<br>[0.50] |
| Full Vacc rate x NorthAm | 0.129<br>[3.68]*** | 0.164<br>[4.43]*** | 0.186<br>[4.59]*** |
| Full Vacc rate x SouthAm | 0.212<br>[5.97]*** | 0.220<br>[5.84]*** | 0.218<br>[5.39]*** |
| Full Vacc rate x Oceania | 0.013<br>[0.30] | 0.076<br>[1.38] | 0.098<br>[1.46] |
| Aged 70+ (percent) | 0.488<br>[7.74]*** | 0.597<br>[7.96]*** | 0.671<br>[8.10]*** |
| Life expectancy | -4.691<br>[-5.78]*** | -5.256<br>[-6.30]*** | -4.918<br>[-5.11]*** |
| Population density | -0.062<br>[-2.96]*** | -0.060<br>[-2.43]** | -0.062<br>[-2.08]** |
| GDP per capita | -0.201<br>[-3.14]*** | -0.121<br>[-1.60] | -0.058<br>[-0.64] |
| Stringency index | 0.395<br>[5.06]*** | 0.373<br>[4.35]*** | 0.407<br>[4.47]*** |
| No. hospital beds | 0.162<br>[3.90]*** | 0.184<br>[3.98]*** | 0.224<br>[4.26]*** |
| Cardiovasc. death rate | 0.369<br>[5.24]*** | 0.501<br>[6.51]*** | 0.642<br>[7.34]*** |
| Diabetes prevalence | 0.703<br>[8.94]*** | 0.719<br>[7.98]*** | 0.810<br>[8.03]*** |
| Constant | 13.433<br>[3.97]*** | 14.285<br>[4.12]*** | 11.204<br>[2.84]*** |
| Observations | 963 | 878 | 800 |
| R-squared | 0.785 | 0.760 | 0.729 |

*Notes:* Dependent variable is defined as logarithm of an end-of-the-week 7-day moving average of the per-million number of new COVID deaths. The lag of full vaccination rate is indicated in the head of the column. All variables are specified in natural logarithms. The model is estimated by Pooled OLS and includes time fixed effects.
Robust t-statistics in brackets, \*\*\* p<0.01, \*\* p<0.05, \* p<0.1.

**Table A7:** Impact of vaccination threshold rate on number of new cases and deaths, all countries

|  | (1) | (2) | (3) | (4) |
| --- | --- | --- | --- | --- |
|  | **Cases** |  | **Deaths** |  |
|  | Lag 3<br>weeks | Lag 4<br>weeks | Lag 3<br>weeks | Lag 4<br>weeks |
| Initial / Lagged No. cases per mill. | 0.595<br>[18.73]*** | 0.589<br>[19.73]*** | 0.796<br>[38.31]*** | 0.763<br>[30.49]*** |
| Reproduction rate_1 | 6.594<br>[15.05]*** | 6.930<br>[16.21]*** |  |  |
| Full Vacc rate | -0.155<br>[-2.51]** | -0.174<br>[-2.82]*** | -0.022<br>[-0.45] | -0.021<br>[-0.32] |
| Full Vacc rate x Vacc (40-70%) | 0.050<br>[1.85]* | 0.064<br>[2.30]** | -0.054<br>[-3.06]*** | -0.049<br>[-2.30]** |
| Full Vacc rate x Vacc (>70%) | -0.082<br>[-2.63]*** | -0.066<br>[-2.06]** | -0.051<br>[-2.28]** | -0.066<br>[-2.48]** |
| Aged 70+ (percent) | 0.551<br>[6.44]*** | 0.580<br>[6.57]*** | 0.692<br>[12.82]*** | 0.824<br>[13.47]*** |
| Life expectancy | -0.673<br>[-0.52] | -0.619<br>[-0.49] | -5.690<br>[-6.89]*** | -6.336<br>[-7.50]*** |
| Population density | 0.002<br>[0.08] | -0.002<br>[-0.08] | -0.119<br>[-7.41]*** | -0.118<br>[-6.39]*** |
| GDP per capita | 0.446<br>[4.79]*** | 0.474<br>[5.01]*** | -0.257<br>[-4.21]*** | -0.150<br>[-2.17]** |
| Stringency index | 0.353<br>[3.43]*** | 0.420<br>[4.36]*** | 0.413<br>[5.70]*** | 0.414<br>[5.14]*** |
| No. hospital beds | 0.014<br>[0.22] | 0.035<br>[0.52] | 0.105<br>[2.47]** | 0.110<br>[2.37]** |
| Cardiovasc. Death rate | 0.482<br>[4.93]*** | 0.557<br>[5.57]*** | 0.145<br>[2.23]** | 0.266<br>[3.67]*** |
| Diabetes prevalence | 0.298<br>[2.98]*** | 0.336<br>[3.28]*** | 0.797<br>[12.23]*** | 0.810<br>[10.87]*** |
| Constant | -9.279<br>[-1.70]* | -10.752<br>[-2.01]** | 19.077<br>[5.56]*** | 20.037<br>[5.71]*** |
| Observations | 1,002 | 914 | 963 | 878 |
| R-squared | 0.668 | 0.683 | 0.769 | 0.742 |

*Notes:* Table contains results estimating the models (1) and (2). Dependent variable is defined as logarithm of an end-of-the-week 7-day moving average of the per-million number of new COVID cases and deaths. The lag of full vaccination rate is indicated in the head of the column. All variables are specified in natural logarithms. The models are estimated by Pooled OLS and include time fixed effects.
Robust t-statistics in brackets, \*\*\* p<0.01, \*\* p<0.05, \* p<0.1.

**Table A8:** Impact of vaccination rate on number of new COVID cases in 2021 compared to 2020, all countries

|  | (1)<br>Lag 2<br>weeks | (2)<br>Lag 3<br>weeks |
| --- | --- | --- |
| Initial No. cases per mill. | 0.456<br>[6.89]*** | 0.413<br>[6.20]*** |
| Reproduction rate_1 | 90.428<br>[3.23]*** | 91.710<br>[2.97]*** |
| Full Vacc rate | -2.204<br>[-0.24] | -1.967<br>[-0.20] |
| Full Vacc rate x Vacc (40-70%) | 18.380<br>[3.31]*** | 18.582<br>[3.06]*** |
| Full Vacc rate x Vacc (>70%) | -12.341<br>[-1.96]* | -11.366<br>[-1.69]* |
| Aged 70+ (percent) | 81.061<br>[5.59]*** | 87.373<br>[5.53]*** |
| Life expectancy | -675.672<br>[-4.09]*** | -811.034<br>[-4.49]*** |
| Population density | -6.756<br>[-1.21] | -9.004<br>[-1.49] |
| GDP per capita | 32.762<br>[2.08]** | 37.130<br>[2.17]** |
| Stringency index | 2.079<br>[4.56]*** | 2.090<br>[4.37]*** |
| No. hospital beds | -6.300<br>[-0.56] | -4.597<br>[-0.38] |
| Cardiovasc. Death rate | 108.446<br>[5.09]*** | 113.676<br>[4.97]*** |
| Diabetes prevalence | 43.550<br>[1.82]* | 50.497<br>[1.93]* |
| Constant | 1,894.905<br>[2.70]*** | 2,396.972<br>[3.12]*** |
| Observations | 968 | 882 |
| R-squared | 0.254 | 0.253 |

*Notes:* Table contains results estimating the model (1). Dependent variable is a difference in per-million number of new COVID cases. The dependent variable, initial / lagged number of cases, reproduction rate and stringency index are defined as a difference in the corresponding variable between the same 16 weeks of August to November in 2021 and 2020. For other variables their 2021 values were taken. The lag of full vaccination rate is indicated in the head of the column. The model is estimated by Pooled OLS and includes time fixed effects.
Robust t-statistics in brackets, \*\*\* p<0.01, \*\* p<0.05, \* p<0.1.

**Table A9:** Impact of vaccination rate on number of hospitalizations, ICU admissions and deaths in 2021 compared to 2020

|  | (1)<br>Lag 3<br>weeks<br>Hospitaliz-<br>ed | (2)<br>Lag 3<br>weeks<br>ICU | (3)<br>Lag 3<br>weeks<br>Deaths | (4)<br>Lag 4<br>weeks<br>Hospitaliz-<br>ed | (5)<br>Lag 4<br>weeks<br>ICU | (6)<br>Lag 4<br>weeks<br>Deaths |
| --- | --- | --- | --- | --- | --- | --- |
| Lagged No. cases per mill. | 0.339<br>[13.97]*** | 0.051<br>[13.31]*** | 0.008<br>[14.51]*** | 0.338<br>[13.66]*** | 0.051<br>[13.68]*** | 0.008<br>[13.75]*** |
| Full Vacc rate | -293.503<br>[-8.09]*** | -39.630<br>[-8.91]*** | -0.348<br>[-2.93]*** | -297.361<br>[-9.48]*** | -41.320<br>[-9.15]*** | -0.343<br>[-2.64]*** |
| Full Vacc rate x Vacc (40-70%) | -12.560<br>[-2.54]** | -1.140<br>[-1.33] | -0.090<br>[-1.52] | -17.538<br>[-3.45]*** | -2.013<br>[-2.43]** | -0.141<br>[-2.21]** |
| Full Vacc rate x Vacc (>70%) | 2.899<br>[0.54] | -0.000<br>[-0.00] | 0.048<br>[0.74] | -1.836<br>[-0.33] | -0.740<br>[-0.76] | 0.021<br>[0.31] |
| Aged 70+ (percent) | 91.163<br>[4.27]*** | 9.467<br>[3.04]*** | 0.929<br>[5.34]*** | 91.068<br>[4.17]*** | 8.086<br>[2.57]** | 1.007<br>[5.20]*** |
| Life expectancy | -813.521<br>[-2.04]** | -37.659<br>[-0.54] | -9.038<br>[-4.12]*** | -983.504<br>[-2.33]** | -6.545<br>[-0.09] | -9.925<br>[-4.25]*** |
| Population density | 7.217<br>[2.02]** | -1.316<br>[-2.66]*** | 0.019<br>[0.37] | 6.875<br>[1.88]* | -1.356<br>[-2.67]*** | 0.031<br>[0.49] |
| GDP per capita | 79.422<br>[3.81]*** | 17.923<br>[5.90]*** | 0.425<br>[2.84]*** | 84.744<br>[3.89]*** | 18.713<br>[5.92]*** | 0.523<br>[3.24]*** |
| Stringency index | 2.632<br>[5.10]*** | 0.153<br>[2.26]** | 0.034<br>[7.66]*** | 2.588<br>[5.01]*** | 0.148<br>[2.24]** | 0.037<br>[7.30]*** |
| No. hospital beds | -89.218<br>[-4.21]*** | -2.431<br>[-0.86] | -0.038<br>[-0.31] | -86.745<br>[-3.93]*** | -1.746<br>[-0.60] | -0.034<br>[-0.26] |
| Cardiovasc. Death rate | 13.374<br>[0.45] | -13.110<br>[-2.87]*** | 1.438<br>[6.11]*** | -3.452<br>[-0.11] | -13.850<br>[-2.87]*** | 1.462<br>[5.73]*** |
| Diabetes prevalence | 27.049<br>[1.74]* | 14.151<br>[6.51]*** | 1.214<br>[5.97]*** | 24.132<br>[1.46] | 14.932<br>[6.61]*** | 1.304<br>[5.42]*** |
| Constant | 3,715.977<br>[2.07]** | 167.915<br>[0.54] | 24.915<br>[2.77]*** | 4,524.510<br>[2.36]** | 37.180<br>[0.11] | 27.400<br>[2.79]*** |
|  | 281<br>0.840 | 252<br>0.797 | 1,006<br>0.567 | 257<br>0.860 | 231<br>0.820 | 918<br>0.554 |

*Notes:* Table contains results estimating the model (2). Dependent variables are differences in per-million number of hospitalizations, ICU admissions and deaths in 2021 relative to 2021. The dependent variable, lagged number of cases and stringency index are defined as a difference in the corresponding variable between the same 16 weeks of August to November in 2021 and 2020. For other variables their 2021 values were taken. The lag of full vaccination rate is indicated in the head of the column. The model is estimated by Pooled OLS and includes time fixed effects.
Robust t-statistics in brackets, \*\*\* p<0.01, \*\* p<0.05, \* p<0.1.

## Footnotes

8 As an example see Singanayagam et al (2021).

9 We consider the latest evidence from United Kindgdom compiled by the UK Health Security Agency published in »COVID-19 vaccine surveillance report (week 45)«. In this report, vaccine effectiveness against symptomatic COVID-19 has been assessed in England based on community testing data linked to vaccination data from the National Immunisation Management System (NIMS), cohort studies such as the COVID Infection Survey and GP electronic health record data. The report focuses mostly on the Delta variant that has been a prevalent COVID-19 variant in the UK and other countries since the early summer of 2021.

10 See COVID-19 vaccine surveillance report, week 45 (UK Health Security Agency, 2021).

11 See full explanation in the Section on data collection.

12 See for example Singanayagam et al (2021).

13 https://github.com/owid/covid-19-data/tree/master/public/data

14 The <u>Oxford Coronavirus Government Response Tracker (OxCGRT)</u> project calculate a Stringency Index, a composite measure of nine of the response metrics measuring strictness of government policies. For details see Hale et al. (2021).

15 The last available data at the time when this research was conducted.

16 Note that these results should not be interpreted as the effectiveness of vaccines against new COVID infections, but as the effectiveness of different vaccination levels across countries in containing the spread of infections.

17 Note that there is no data available for number of hospitalized and ICU patients for countries in South America and Oceania.

18 The period of comparison is 16 weeks between August 1 and November 14, 2021 and 16 weeks between August 2 and November 15, 2020.

